# The role of interleukin-10 in immune response to hepatitis B virus and liver cancer co-existence dynamics

**DOI:** 10.1101/2024.07.14.24310388

**Authors:** Paul Chataa, Samuel M. Naandam, Francis T. Djankpa

## Abstract

Experimental evidence confirms that interleukin-10 plays a critical role in clearing acute hepatitis B virus infection. This paper aims to develops a mathematical model to explore the dynamics of how the immune system responds to hepatitis B virus (HBV) and coexisting liver cancer within the liver cell population. Unlike previous models; we categorize liver cells into various stages of infection. We determine the invasion probability for transmission dynamics, specifically the basic reproduction number, ℝ_0_, for populations of uninfected macrophages with and without cancer cells. Stability analyses of virus-free and virus equilibrium states are provided, along with numerical simulations to validate analytical findings. The impact of different branches of the immune response on model dynamics is assessed. Simulations predict the time at which T helper-1 cells surpass cytotoxic T cells (switching time), correlating positively with the proliferation rate of interleukin-10 (*ρ*_3_). Further numerical simulations demonstrate that interleukin-10 contributes to HBV persistence by inhibiting the immune response, thereby allowing the virus to evade immune surveillance and establish chronic infection through the suppression of cytotoxic T lymphocytes (CTLs), which are essential for clearing infected cells.

## 1 Introduction

A viral infection that targets the liver cells called hepatitis B can result in both acute and chronic illnesses. According to [1], this condition kills 750, 000 people per year, roughly 300, 000 of which result in liver cirrhosis and hepatocellular carcinoma [2]. A third of the world’s population is thought to be infected with the hepatitis B virus, making it not only one of the oldest but also one of the most dangerous viral dangers to human health [3]. According to the World Health Organisation (WHO) estimates, 1.5 million new cases of hepatitis B are reported annually, leaving 296 million people living with the virus. WHO estimates again indicated that hepatitis B caused 820000 in fatalities in 2019, with the majority being caused by cirrhosis and hepatocellular carcinoma, the predominant form of liver cancer. Every year, millions of individuals worldwide perish from liver cancer, and current trends suggest that millions more people will do so in the future. Reports from Europe and America shows that the hepatitis B infection is quite low (i.e., less than 1%). With a chronic infection incidence of 5 to 10 per 100 adults in Asia and Africa, the disease is still a significant burden in these regions.

The hepatitis B virus, a member of the hepadnaviridae family of viruses, is what causes hepatitis B disease. Once a person is infected, it is particularly challenging to get rid of the hepatitis B virus due to its largely double-stranded structure [2]. The incubation periods for the hepatitis B virus range from 30 to 180 days. The virus may be detected within 30–60 days of infection, and if it is transferred during infancy or childhood, it may persist and result in chronic hepatitis B. The virus is frequently vertically transferred from mother to child, causing recurrent infection and, in the majority of instances (approximately 90%), chronic infection [4, 5]. The second potential route for virus spread is through horizontal adult-to-adult sexual contact, intravenous drug use, unhygienic behavior, and blood transfusion. Only 5-10% of adults have persistent infections as a result of this sort of transmission, according to [4] and [5].

When the body generates sufficient immune responses against the infection, these adults recover. Activation of robust and diversified CD4 (T helper 1) and CD8 (cytotoxic T lymphocytes) T-cells, production of protective, neutralizing antibodies against HBV surface antigen (HBsAg), and expression of antiviral cytokines in the liver, such as gamma interferon (type-2 interferons) and tumor necrosis factor alpha [6, 5], and the creation of refractory cells, which are immune to reinfection [7, 8] are examples of such responses. However, newborns who are not immunized and immunosuppressed adults tend to move to chronic HBV infection stage [9]. These people have humoral and cellular immune systems that are weak and ineffective, which leads to ongoing viral replication and HBV surface antigenemia [2, 10]. The relative contributions of the immune system’s many components are poorly understood, particularly the functions of anti-inflammatory cytokines like interleukin-10 in the development and progression of infection.

The World Health Organization has set a goal of eliminating liver cancer and the hepatitis B virus (HBV) by 2030 [11, 12]. Treatment of sick people as well as immunization of newborns and susceptible adults with the hepatitis B vaccine are now used as control techniques to lessen the spread of the hepatitis B virus. Campaigns aimed at educating people about how alcohol and smoking can prevent liver cancer and help slow the spread of the illness is another way to control the spread of these two diseases. These preventative measures are meant to stop acute hepatitis B virus infection and prevent people with liver problems from developing liver cirrhosis again. However, the incidence of hepatitis B and liver cancer is alarmingly high, with millions of cases and deaths each year. Although there has been progress toward eliminating HBV, the disease still poses a serious threat to public health. 316 million persons worldwide were living with chronic HBV infection in 2019 [13]. This represents a prevalence of chronic HBV infection of 4.1%.

The likelihood of clearing liver cancer is decreased when HBV infection coexist with liver cancer. Therefore, it is essential for the research community to conduct scientific study on hepatitis B and liver cancer in both clinical and theoretical forms. The development of effective hepatitis B and liver cancer therapies is a vast area of medical study that will influence our comprehension of tumor-immune dynamics. Although the majority of people (about 90% in adults) assemble a successful and defensive cell-driven mechanism that prevents them from developing chronic hepatitis B disease, which will eventually result in liver cancer, hepatitis B is still a leading cause of death globally (about 3 million deaths per year), according to estimates [2].

Numerous researchers have examined different facets of HBV and liver cancer dynamics and the immune response during infection using mathematical models. In order to investigate acute HBV infection and the significance of time lag in effector cell activation and expansion, [3, 14] expanded a standard model of immune response to incorporate the time delay in recruit naive T cells. Subsequently, they also investigated the function of pre-existing or vaccine-induced antibodies in containing the HBV infection [1]. Instead of using a mass action to account for a finite liver size and susceptibility to HBV infection, [15] employed a typical incidence function in their study of HBV transmission dynamics. A time-delayed version of the model put forth in [15] has been created by [16]. [17] model was able to more accurately describe the existing data and produce more realistic results for the basic reproduction number by using a standard incidence and a logistic growth for the hepatocyte population. [18] have examined potential inadequacies in the synchronization of distinct branches of the adaptive immune response, particularly the CTLs and antibodies, in the context of HBV infection. Yet, current modeling studies have not been able to aid in the development of successful HBV total eradication and drug treatments. A reason for this may lie in the fact that almost all models of HBV and liver cancer ignore the interleukin-10 and the differentiation of naive T cells aspects of infection which we have determined to be quite important for designing HBV and liver cancer control strategies.

Moreover, it is impossible to ignore the critical part the immune system plays in the dynamics of the hepatitis B virus and liver cancer infection. Interleukin-10 (*I*_10_) is a cytokine that modulates both innate and adaptive immunity, primarily by exerting anti-inflammatory effects. So, formulating a new model to examine how interleukin-10 and other components of the immune system regulates both cell-mediated (most especially the production of T helper-1 and cytotoxic T cells) and innate immunity proved beneficial. Thus, we concentrated on limiting the concentration of the virus and the damage to the liver. We achieved this goal using different strategies which include interferon immunity (i.e type-1 and type-2) by removing the substrate that the virus needs for reproduction (i.e., the healthy cells), cellular immunity (T cells, NK cells, effector B cells, interleukin-10 cells) by removing the source of new viruses (i.e., the infected cells), and adaptive immunity (HBV-antibodies) by lowering the effectivity concentration of the virus. Another innovative part of our model is how it predict when T helper-1 (*T*_1_) cells will outnumber cytotoxic T cells (*T*_2_), a period we call the “switching time”. This period predict when acute hepatitis B infection transitions to chronic infection and eventually to liver cancer and the conditions that led to these transitions.

The paper is organized as follows; Section 2 is devoted to the mathematical formulation of the model. The model basic properties was presented in Section 3. In Section 4 we present the model analysis. Local asymptotic stability of the virus-free equilibrium and global asymptotic stability of virus-free analysis was presented in Section 5. In Section 6, we study the numerical results and sensitivity analysis of the proposed models and present the results in the form of plots, and a discussion of the results is presented in section 7.

## 2 Materials and methods

### 2.1 Basic Model Formulation

A compartmental mathematical model for the immune response dynamics to hepatitis B virus and liver cancer infection was constructed in order to comprehend the numerous changes that occur in the immune system’s response to the coexistence dynamics of these two diseases. The model is based on information obtained from new scientific explanations of the basic features of infections with the hepatitis B virus and infections that cause liver cancer, as well as on theoretical and experimental studies conducted by other researchers [5, 19, 14, 12]. The outcome of a hepatitis B virus infection and the rate at which acute infections lead to liver cancer are significantly influenced by the roles of antibodies, cell-mediated immune responses, and innate immune system responses as well as the cytokines. The total number of human liver cells, or hepatocytes, was categorized into two populations: uninfected macrophages without cancer *M*_0_, and those with cancer *M*_1_. Both uninfected macrophages with and without cancer (i.e *M*_0_ and *M*_1_) are triggered from the blood stream to the site of infection because the hepatitis B virus is present in the body cells. Both of the uninfected macrophages then phagocytose the viruses, which causes them to become infected at time *t*. These infected macrophages are identified by *I*_*M*_. It is assumed that both uninfected macrophages are created at constant rates 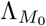 and 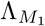 and die at constant rates 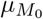 and 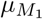 per cell. Hepatitis B virus particles infect them at a rate proportional to the product of *M*_0_, *M*_1_, and *V* (i.e., *β*_1_*V M*_0_ and *β*_2_*V M*_1_) with constant proportionality rates of *β*_1_ and *β*_2_, respectively. Infected macrophages pass away at a steady rate of 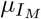 per cell. In the class of infected macrophages, some of the individual macrophages develop cirrhosis, pick up liver cancer, and proceed at a rate of *ζ* to the liver cancer macrophage population. Through contaminated blood transfusions and having sex with an infected individual, liver cancer macrophages can get hepatitis. Hepatitis B virions *V* are recruited at a rate of Λ_*V*_ whereas infected macrophages create additional free virions at a rate of *ω*. The virus degenerates at a constant rate of *µ*_*V*_ in each cell. Figure 1 shows interactions between all cell types in the human liver.

**Figure 1:**
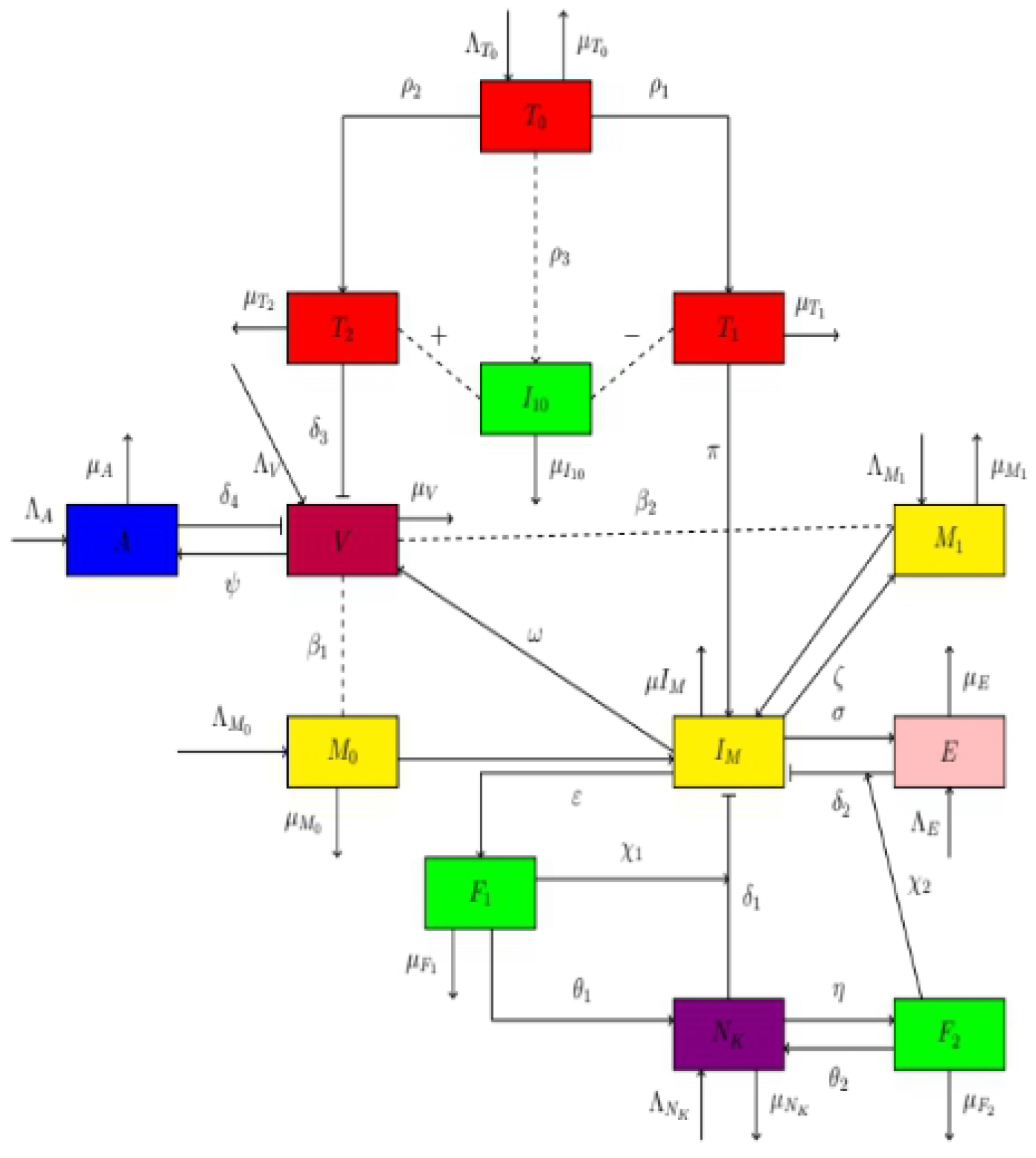
Show the schematic diagram of immune response to HBV and liver cancer co-existence. Yellow rectangles indicate host liver cells (uninfected macrophages with and without cancer, and infected macrophages) population, red rectangles denote T cell’s population, blue rectangle represent antibodies population, pink rectangle represent effector B cells population, green rectangles show cytokines (type-1 and type-2 interferon) population, the violet rectangle is the innate (*N*_*K*_ cells) population, and purple rectangle indicates virus particles (virions) population. Also, thick lines with arrowheads represent contacts, thick lines with bar heads represent clearance (destroy) while dash lines represent interactions.

Invading infections are specially recognized by antibodies, which bind to them and render them ineffective. Sub-viral particles (SVPs), which can occur in quantities up to a thousand to one million times higher than the infectious virions, are produced in excess by HBV-infected cells during infections. These SVPs may have an impact on how the immune system of the host responds to the HBV infection. According to [1, 20], the sub-viral particles serve as immune system spies that tempt antiviral antibodies away from binding to the hepatitis B virus. They may also promote tolerance during neonatal infection, delaying the development of neutralizing antibodies and allowing the hepatitis B virus to avoid antibody detection. The creation of macrophages, T cells, and cytokines triggers a unique immune response that frequently mediates this issue. We assume that in the absence of infection, HBsAg-specific antibodies *A* are created over time rate Λ_*A*_ and degrade at a per capita rate *µ*_*A*_. Nevertheless, throughout an infection, antibodies are created at a rate *ψ* that is proportionate to the viral load and kill virions at a rate of *δ*_4_ [21]. Macrophages T cells are created at a rate of 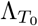 and perish at a constant rate of 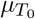 in each cell. Following internalization by the infected macrophages, the virus continues to spread by feigning an acute stage (i.e., the stage at which virus reproduction is extremely small) for a predetermined amount of time in order to evade intracellular killing systems. The virus population in the host liver cells grows as a result of the virus’s ongoing intracellular replication in an infected macrophages cell. Naive T cells from the bone marrow’s thymus are induced, triggered, and activated as a result of virus replication inside infected macrophages up to a point. The pro-inflammatory cytokines actions of the virus cause the naive T cells to differentiate into either T helper 1; *T*_1_ or T helper 2; *T*_2_ cells. The number of infected macrophages per unit area and the amount of virus present, respectively, determine the cellular *T*_1_ and *T*_2_ immunological responses to HBV infection. While T helper 1 cells *T*_1_ differentiate at a rate *ρ*_1_ and die at a per capita constant rate 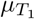 with each cell specialized in carrying out a specific task, T helper 2 cells become cytotoxic T lymphocytes *T*_2_ at a differentiation rate *ρ*_2_ and die at a per capita constant rate 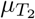. While T helper 1 cells; *T*_1_ activate the infected macrophages cells to generate additional cytokines at a rate of *π*, cytotoxic T cells destroy their targeted virus cells primarily by releasing cytotoxic granules to kill the virus cell at a rate of *δ*_3_.

The naive T cells undergo additional differentiation to form regulatory T cells, which control the generation of *T*_1_ and *T*_2_ cells. It has been discovered that the production of regulatory T cells, which have been found to perform a lytic activity for the production of interleukin-10; *I*_10_, is delayed as a result of the virus’s ability to appear temporarily inactive during the acute stage in the infected macrophages [22]. Interleukin-10 is a cytokine that predominantly has anti-inflammatory effects and affects both *T*_1_ and *T*_2_ adaptive immunity. During HBV infections, it also coordinates the innate immune system and antibodies. Naive T cells create *I*_10_ at a rate of *ρ*_3_ and it dies at a constant rate of 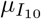 in each cell. It alternately suppresses and supports the development of *T*_1_ and *T*_2_ cells. Antigen-presenting cells (APCs) cannot produce pro-inflammatory cytokines like *I*_12_, and they are also prevented from up-regulating molecules involved in antigen presentation and lymphocyte activation (i.e. the minus sign stands for down regulation of cytokines), which is how *I*_10_ suppresses T helper 1 cells. Although these cytokines primarily have suppressive effects on the immune system, they also have certain immune-stimulating properties, such as increasing the generation of cytotoxic T lymphocytes also known as T helper 2 cells. The plus sign denotes cytokine up-regulation.

Another method by which the immune system can combat and stop the progression of HBV infection into liver cancer is through effector B cells, also referred to as plasma cells. These effector cells releases antibodies, heals infected cells, and primes T cells (including cytotoxic T cells and T helper 1 cells), which trigger cell-mediated reactions. Effector cell levels are maintained at a certain homeostatic level after viral clearance as a result of long-lived plasma and memory B cells [6]. When viral peptide MHC class I molecules, which include viral proteins, are displayed on the cell surface, effector B lymphocytes begin to target those cells as infected cells. In the absence of infection, we assumed that effector B cells proliferate at a rate of Λ_*E*_ and perish at a constant rate of *µ*_*E*_ per cell. We model these two impacts; killing and curing of infected cells in one reaction, where effector B cells kill and cure infected macrophages at a rate *δ*_2_. This is because it has been demonstrated that cured cells lose their resistance to productive infection at a slow rate up to the order of 10^5^ per day, and effector cells also kill infected macrophage cells. When there is an infection, the population of effector cells increases by a factor of *σI*_*M*_ *E*, where *σ* is the maximum proliferation rate. This happens in a contaminated cell density-dependent manner which depends on the density of an antigen.

The hepatitis B virus is highly adaptable and has developed ways of stopping MHC molecules from getting to the cell surface to display viral peptides to avoid being detected by T cells. If this happens, the T cell may not know that there is a virus inside the infected cell. However, another immune cell specializing in killing cells with a reduced number of MHC class I molecules on their surface is the natural killer cell or *N*_*K*_ cell for short. When the *N*_*K*_ cells finds a cell displaying fewer than normal MHC molecules it releases toxic substances, in a similar way to cytotoxic T cells to kill the viral-infected cell. Similar to effector B cells, in the absence of infection, natural killer *N*_*K*_ cells, which are the first line of defense against non-self pathogens are considered to grow at a rate 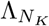, and die at a per capita constant rate 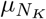. When examining the immune response to HBV and liver cancer infection co-dynamics, the importance of cytokines in immune system dynamics cannot be understated. Interferon, a class of tiny proteins produced and released by virally infected macrophage cells, is crucial for immunological defense against hepatitis B virus infection. By directly impeding the ability of the virus to replicate within an infected macrophages cell, interferon stops viral replication. We refer to the Type-1 interferon (*IFN*_*α/β*_) as *F*_1_, which is created by infected macrophages cells at a rate of *ε* and destroyed at a per-cell rate of 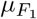 [2, 8]. Type-2 interferon *IFN*_*γ*_, on the other hand, is denoted by *F*_2_ which is also produced by natural killer cells *N*_*K*_ [6, 9, 10] at rate *η* and they are lost at a per capita rate 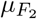.

*IFN*_*γ*_ generates protein-10 that can both energize and enroll *N*_*K*_ cells [10], whereas *IFN*_*α/β*_ are able to activate the natural killer *N*_*K*_ cells during infection [23]. Therefore, it is assumed that the cumulative action of interferon on initiating *N*_*K*_ cells happens at *θ*_1_*N*_*K*_*F*_1_ + *θ*_2_*N*_*K*_*F*_2_; where *θ*_1_ represents the rate at which Type-1 interferon activates *N*_*K*_ cells and *θ*_2_ represents the rate at which Type-2 interferon activates *N*_*K*_ cells respectively. In addition to increasing the number of new *N*_*K*_ cells produced, *IFN*_*α/β*_ and *IFN*_*γ*_ boost the the *N*_*K*_ cells’ pathogenicity and effector B cells, respectively [24]. Hence, we assumed that natural killer cells and effector B cells destroy infected macrophages cells at *δ*_1_ (1 + χ_1_*F*_1_) *I*_*M*_ *N*_*K*_ and *δ*_2_ (1 + χ _2_*F*_2_) *I*_*M*_ *E*; where χ_1_ represent the rate at which Type-1 interferon increase the pathogenicity of *N*_*K*_ cells and χ_2_ represent the rate at which Type-2 interferon increase the pathogenicity of effector B cells respectively. We additionally assume the following in addition to the presumptions listed above:

- It is assumed that a portion of the liver cells have acquired malignancy from external sources such as excessive alcohol intake, smoking etc. We now introduce HBV infection and studied the new co-existence dynamics.
- Time delays in the replication of individual cell components are not taken into account.
- The populations of macrophages cells and virus are assumed to be uniformly distributed over the system at all times.

The general process of the model is described in Figure 1

### 2.2 Model Equations

The following set of differential equations represents the immune response to hepatitis B virus and liver cancer co-existence model, based on the assumptions stated above. Clonal selection theory, mass-action kinetics, and the balances between cell and molecular population proliferation and mortality underpin interactions in the model system.

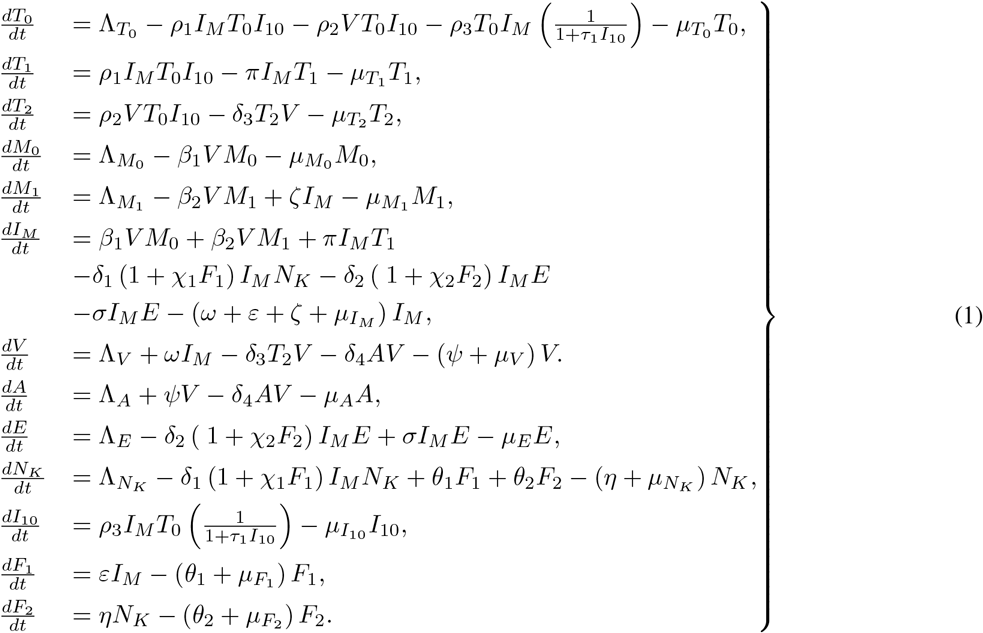

The system described in reference 1 is not dimensionless. The term “non-dimensionless” implies that, in the absence of knowledge regarding the actual values of the variable, we assume these values to be proportional.

## 3 Basic Model Properties

In this section, we present the positivity and boundedness of solutions of the model system.

### 3.1 Positivity of Solutions

To ensure the mathematical and epidemiological relevance of system 1, it is essential to ensure that solutions originating from positive initial conditions remain positive for all *t >* 0. Therefore, it is necessary to establish that each state variable is non-negative, as negative cell population densities are illogical. This outcome is achieved through the lemma provided below.

#### Lemma 1.

If *T*_0_(0) *>* 0, *T*_1_(0) *>* 0, *T*_2_(0) *>* 0, *M*_0_(0) *>* 0, *M*_1_(0) *>* 0, *I*_*M*_ (0) *>* 0, *V* (0) *>* 0, *A*(0) *>* 0, *E*(0) *>* 0, *N*_*K*_(0) *>* 0, *I*_10_(0) *>* 0, *F*_1_(0) *>* 0 and *F*_2_(0) *>* 0, then the solutions *T*_0_(*t*) *>* 0, *T*_1_(*t*) *>* 0, *T*_2_(*t*) *>* 0, *M*_0_(*t*) *>* 0, *M*_1_(*t*) *>* 0, *I*_*M*_ (*t*) *>* 0, *V* (*t*) *>* 0, *A*(*t*) *>* 0, *E*(*t*) *>* 0, *N*_*K*_(*t*) *>* 0, *I*_10_(*t*) *>* 0, *F*_1_(*t*) *>* 0 and *F*_2_(*t*) *>* 0 of model 1 are positive for all *t* ≥ 0.

*Proof*. We define

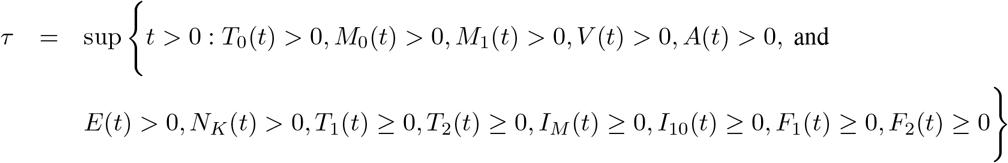

this implies that

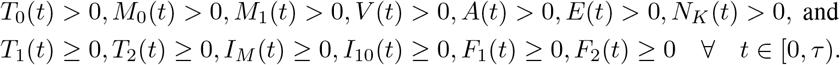

Considering the first equation in system 1, we have

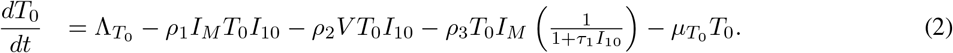

It follows from (2) that

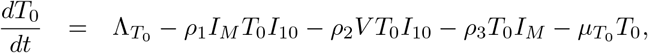

Since 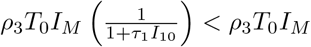. Hence,

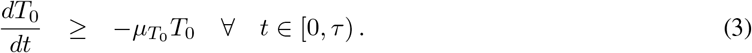

Seperating variables and integrating both sides of (3) from 0 to *τ* gives

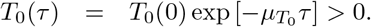

Similarly the remaining equations gives

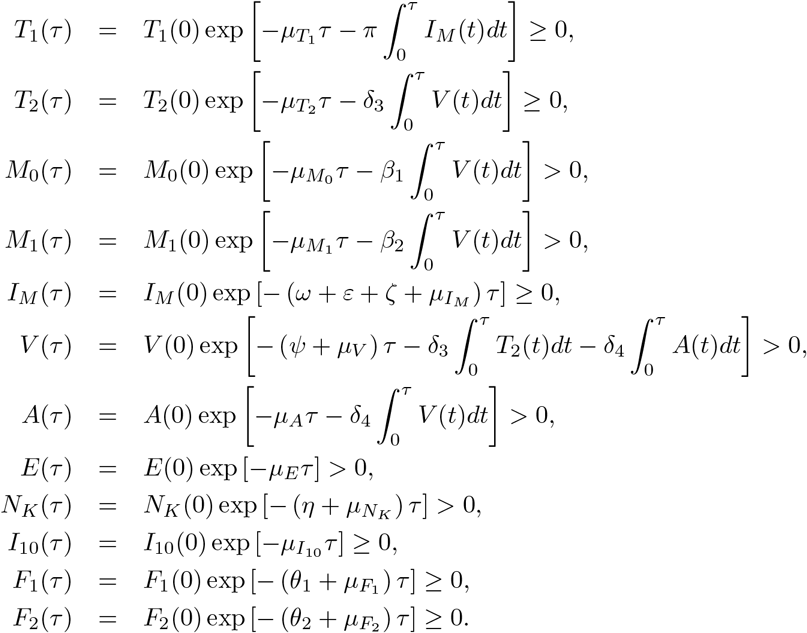

Clearly, we have shown that *T*_0_(*t*) *>* 0, *M*_0_(*t*) *>* 0, *M*_1_(*t*) *>* 0, *V* (*t*) *>* 0, *A*(*t*) *>* 0, *E*(*t*) *>* 0, *N*_*K*_(*t*) *>* 0, and *T*_1_(*t*) ≥ 0, *T*_2_(*t*) ≥ 0, *I*_*M*_ (*t*) ≥ 0, *I*_10_(*t*) ≥ 0, *F*_1_(*t*) ≥ 0, *F*_2_(*t*) ≥ 0. Therefore, the solutions *T*_0_(*t*), *T*_1_(*t*), *T*_2_(*t*), *M*_0_(*t*), *M*_1_(*t*) *>* 0, *I*_*M*_ (*t*), *V* (*t*), *A*(*t*), *E*(*t*), *N*_*K*_(*t*), *I*_10_(*t*), *F*_1_(*t*) and *F*_2_(*t*) of the system 1 remains positive for all *t* ≥ 0. □

### 3.2 Boundedness of Solutions

The model population variables and parameters changes in system 1 were studied keenly under certain conditions. Considering the variables and parameters in the model system to be positive entities for all time values *t* ≥ 0, we show that all possible solutions are uniformly bounded.

#### Lemma 2.

The solutions of the system with any non-negative initial conditions are bounded for all *t* ≥ 0 in the biologically feasible region defines by the set

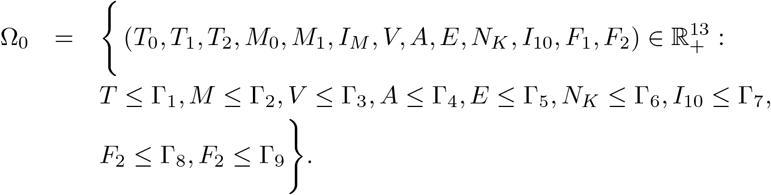

*Proof*. Considering the various T cells (i.e *T*_0_, *T*_1_ and *T*_2_) and summing them to obtain the total T cells population, we have

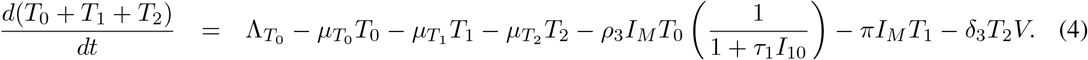

It follows from (4) that

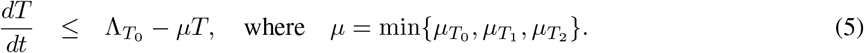

Integrating (5) from 0 to *t*, we obtain

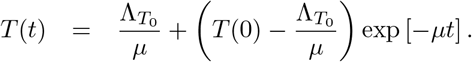

Taking the limit of *T* at *t* → ∞ such that

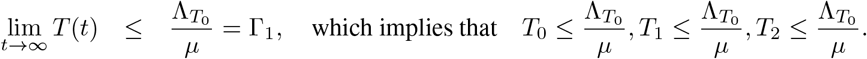

Hence, the T cells population is bounded. That is

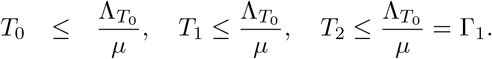

Similarly, the remaining populations give the following

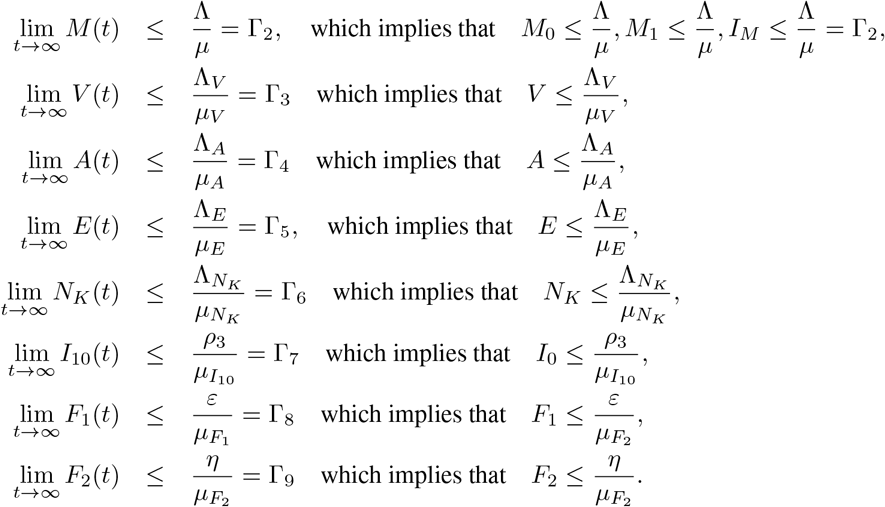

We can therefore conclude that for *t >* 0, any solution of the system is bounded in the region Ω_0_. Thus, it is feasible to consider the dynamics and flow of the T cells, macrophages, virus, antibodies, effector B cells, natural killers and cytokines populations as described by the model system within the invariant region Ω_0_.

## 4 Model Analysis

In this section, we determine the virus-free and virus-persistence equilibria of the model system.

### 4.1 Virus-free Equilibrium and Reproduction Number

It is believed that some liver cells have developed cancer due to external factors like excessive alcohol consumption and smoking. We have now introduced an HBV infection and examined the resulting co-existence dynamics. Hence, at the virus-free equilibrium state, no hepatitis B virus is present which implies that there are no infected macrophages, since both uninfected macrophages (with and without cancer) would have no virus to swallow-up to cause them to be infected. That is *V* ^⋆^ = 0 which immediately implies 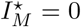 and 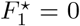. The lack of infected macrophages indicates that the hepatitis B disease is not being transmitted, which in turn means that naive T cells will not perform any lytic activity. Therefore, at the virus-free equilibrium, the populations of all cells involved in the immune interactions are given by

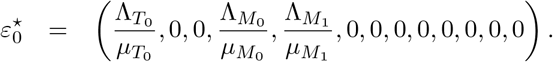

Next, we use the Next Generation Matrix technique described by [25] and use in [26] to calculate the reproduction number of the model system. The number of further HBV and liver cancer infections brought on by a single infected macrophage placed into a population of liver cells that are completely susceptible to infection is known as the reproduction number (ℝ_0_), for an infection. To put it another way, when all liver cells are free of infection, ℝ_0_ represents the amount of secondary infections that one infected macrophage cell produces. The model system’s equations describing the generation of fresh infected macrophages and modifications in their states are the first things we look at. The source of these equations is given by

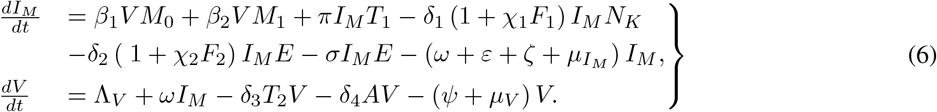

The set of these equations in system 6 is called infected subsystem. Linearizing the infected subsystem about the virus-free steady state is the initial stage, according to the existing rule. When we set

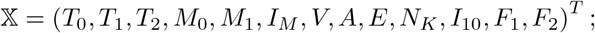

where *T* denote the transpose, then the infected subsection can be written in the form:

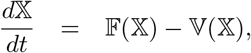

where

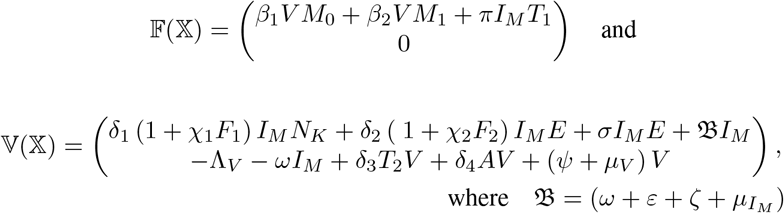

Taking the Jacobian of 𝔽 (𝕏) at the virus-free equilibrium state, we have

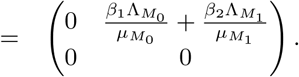

Also taking the Jacobian of 𝕍(𝕏) at the virus-free equilibrium, we obtain

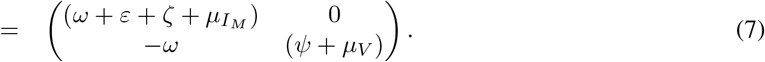

It follows from (7) that

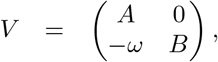

where

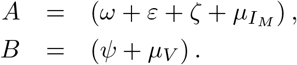

Thus,

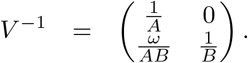

Thus, the next generation matrix which represents the number of secondary infections produced by one infected macrophages cell when all liver cells are uninfected is given as

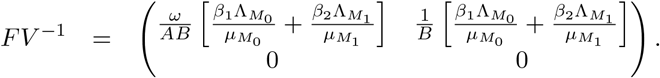

Therefore,

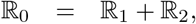

where

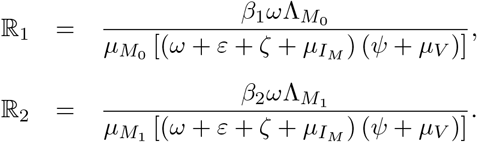

## 5 Model Stability Analysis

The local and global stability analysis of the virus-free equilibrium point, *ε*_0_, as well as the virus-persistence equilibria were investigated in this section.

### 5.1 Local Stability of Virus-free Equilibrium

By calculating the eigenvalues of the linearized Jacobian matrix at the virus-free equilibrium, we are able to ascertain the local stability of the equilibrium when there is a slight disruption. That is when a small number of infected cells are injected into the population of susceptible liver cells, it is possible to completely eliminate the virus infection from the liver cells due to the stability of virus-free equilibrium when ℝ_0_ *<* 1.

#### Theorem 1.

The virus-free equilibrium of system 1 is locally asymptotically stable if ℝ_0_ *<* 1 and unstable if ℝ_0_ *>* 1.

*Proof*. We investigate the proof of Theorem 1 by linearization approach. We make used of the Jacobian matrix associated with system 1 at the virus-free equilibrium and obtain

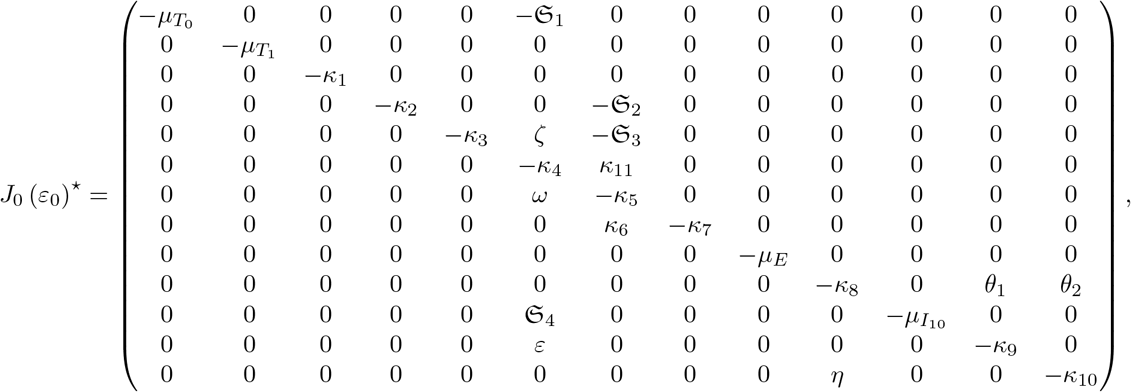

where 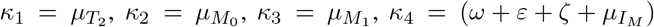, *κ*_5_ = (*ψ* + *µ*_*V*_), *κ*_6_ = *ψ, κ*_7_ = *µ*_*A*_, 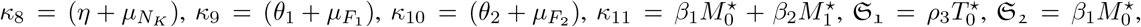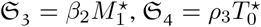

The eigenvaules from the jacobian matrix 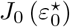 are obtained to be 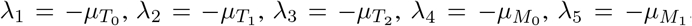, *λ*_6_ = −*µ*_*A*_, *λ*_7_ = −*µ*_*E*_, 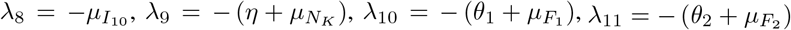 with a polynomial equation

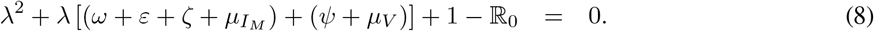

Since all the coefficients of the characteristics polynomial in (8) are positive when ℝ_0_ *<* 1, by the Routh-Hurwitz criterion the solutions to the characteristic polynomial have negative real parts. Therefore all the eigenvalues of the Jacobian matrix 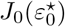 have negative real part when ℝ_0_ *<* 1. Hence, we can conclude based on Routh-Hurwitz criterion that the virus-free equilibrium 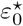 of system 1 is locally asymptotically stable.

### 5.2 Global Stability of Virus-free Equilibrium

#### Theorem 2.

From system 1, the virus-free equilibrium

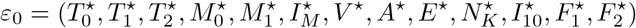

is globally asymptotically stable if ℝ_0_ *<* 1 and conditions (*G*_1_) and (*G*_2_) are satisfied. We apply the approach of [27], to prove the global stability of the virus-free equilibrium.

#### Theorem 3.

If a model system can be written in the form:

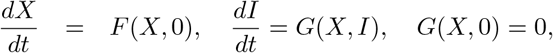

where *X* ∈ ℝ^*m*^ denotes the number of uninfected macrophages cells and *I* ∈ ℝ^*n*^ denotes the number of infected macrophages cells including latent, acute and exposed cells. *U* (*X*^⋆^, 0) denotes the virus-free equilibrium of the system. Then the conditions (*G*_1_) and (*G*_2_) must be satisfied to guarantee local asymptotic stability.

*G*_1_ : 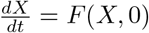, *X*^⋆^ is globally asymptotically stable.

*G*_2_ : *G*(*X, I*) = *AI* − *Ĝ* (*X*, 0) ≥ 0 for (*X, I*) ∈ Ω, where *A* = *D*_*i*_*G*(*X*^⋆^, 0) is a Metzler matrix (the off diagonal elements of *A* are non-negative) and Ω is the region where the model makes biological sense and mathematically well posed. Then the fixed point *U*_0_ = (*X*^⋆^, 0) is globally asymptotically stable equilibrium of the immune response to hepatitis B virus and liver cancer infection model 1 provided ℝ_0_ *<* 1.

*Proof*. From the model system in 1, we have

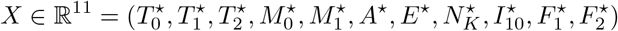

and *I* ∈ ℝ^2^ = (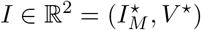, *V* ^⋆^). Hence, for condition (*G*_1_), we have

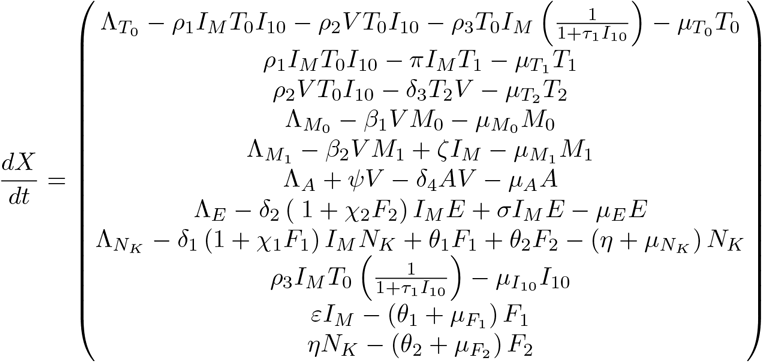

and

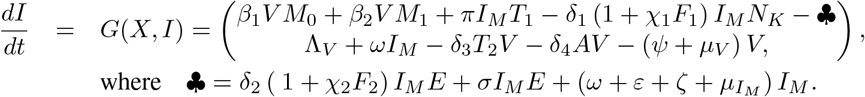

It follows that

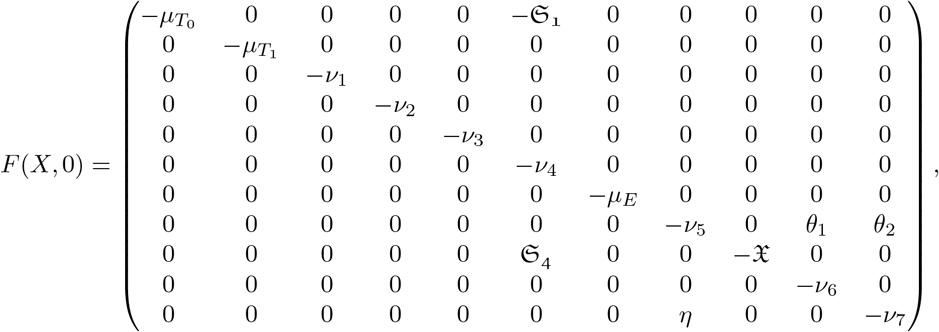

where 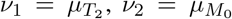, *ν*_4_ = *µ*_*A*_, 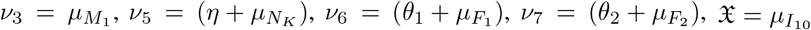.

The eigenvaules from the matrix *F* (*X*, 0) are obtained to be 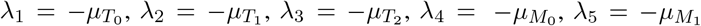, *λ*_6_ = −*µ*_*A*_, *λ*_7_ = −*µ*_*E*_, 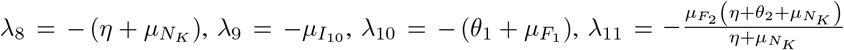

Since all the eigenvalues are real and negative, it follows that *X*^⋆^ is always globally asymptotically stable.

Also, applying Theorem 3 to the immune response model system 1 gives

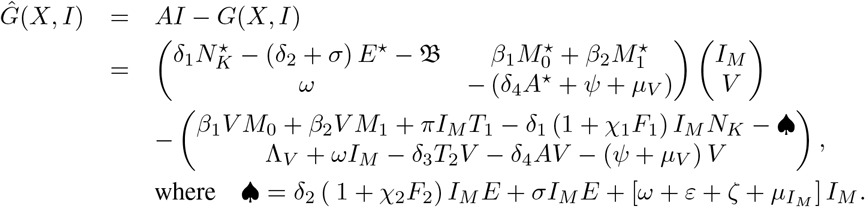

Hence,

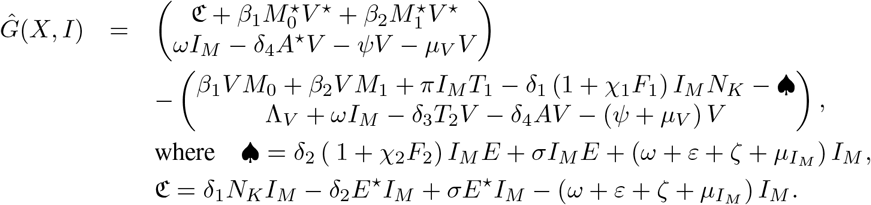

Therefore,

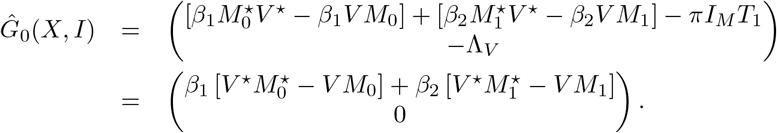

So, *A* is a Metzler matrix with non-negative off-diagonal elements. We observed that

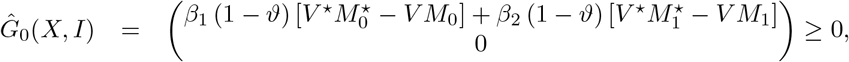

because 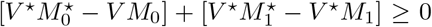. That is in the absence of infection, the proportional product of the macrophages with and without cancer at the virus-free equilibrium are obtained to be 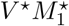 and 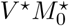 respectively while *V M*_1_ and *V M*_0_ represent the proportion to the product of macrophages with and without cancer in the population. Therefore, the virus-free equilibrium *ε*_0_ is globally asymptotically stable.

### 5.3 Virus-persistence Equilibrium (VPE) and Stability Analysis

In this section, we determine the virus-persistence equilibrium points by solving system 1 simultaneously for the state variables 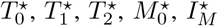, *V* ^⋆^, *A*^⋆^, *E*^⋆^, 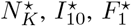 and 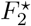 respectively. The virus-persistence equilibrium points are the steady state solutions where the hepatitis B virus and liver cancer infection cannot be totally eradicated but remains to invade the total cells populations. It is assumed that a portion of the liver cells have acquired liver cancer from external sources such as excessive alcohol intake, smoking etc. We now introduce HBV infection and studied the new co-existence dynamics. For the hepatitis B virus infection to remain in the host liver cells, the virus compartment does not turn to zero. As a result of this, the system depends on all the state variables. At the virus present state, uninfected macrophages become infected and this triggers activation of infected macrophages by cytokines. So, at the virus-persistence equilibrium the following equations are satisfied:

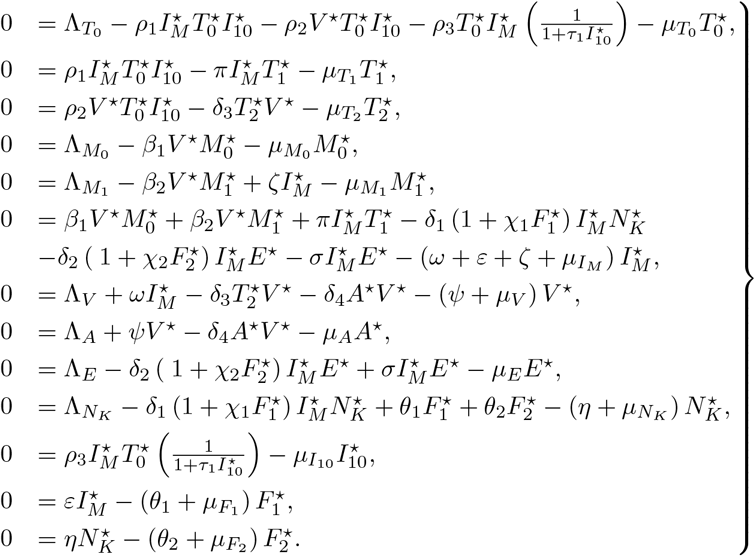

Due to the high dimensionality of system 1, it is prudent to look at a significant number of possible boundary steady states. Hence, in order to systematically find and analyze these steady state, we begin with boundary steady states characterized by the presence hepatitis B virus particles (i.e *V ≠* 0) with different combinations of 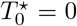 or 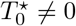, *E*^⋆^ = 0 or *E*^⋆^ *≠* 0 and 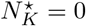or 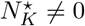.

Case I: For the first scenario where *V* ^⋆^ *≠* 0, 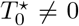, *E*^⋆^ *≠* 0, 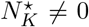, we obtain a boundary steady state of the form:

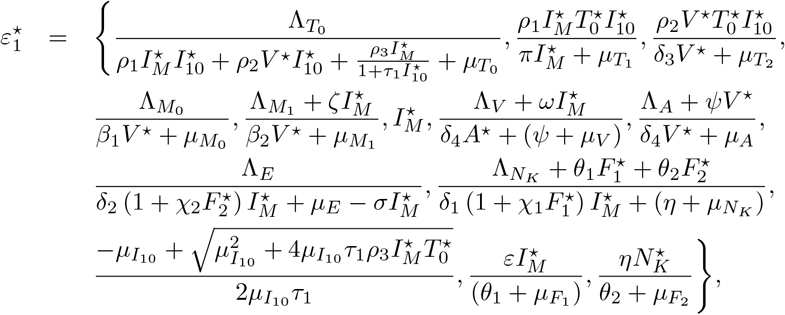

where 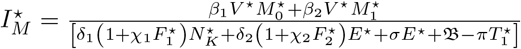 and 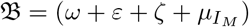.

This equilibrium state 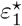 exists when 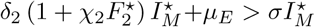, and 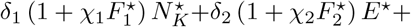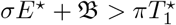. Using the the parameter values in Table 1, we compute the components of 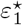 and obtain

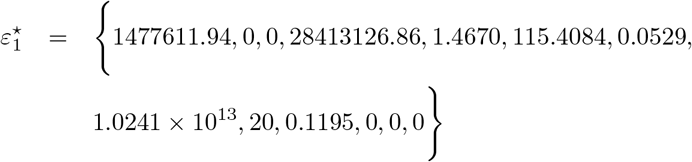

**Table 1:** Parameters of model and their description.

| Parameter | Values | Source | Parameter | Values | Source |
| --- | --- | --- | --- | --- | --- |
| $\Lambda_{T_0}$ | 0.99 cell/day | [28] | $\rho_1$ | $2.9 \times 10^{-4}$ cell/day | [28] |
| $\Lambda_{M_0}$ | $4 \times 10^5$ cell/day | [3, 29, 30] | $\rho_2$ | 0.02 cell/ ml/day | [31] |
| $\Lambda_{M_1}$ | 0.015 cell/day | [32] | $\rho_3$ | $2.03 \times 10^{-7}$ cell/day | [28] |
| $\Lambda_V$ | 0.3 cell/day | [28] | $\delta_1$ | 0.67 cell/day | [1, 21] |
| $\Lambda_A$ | $3.4 \times 10^{12}$ cell/day | [33] | $\delta_2$ | 0.67 cell/day | [1, 21] |
| $\Lambda_E$ | 10 cell/day | [14] | $\delta_3$ | 0.67 cell/day | [1, 21] |
| $\Lambda_{N_K}$ | 0.057 cell/day | [34, 35] | $\delta_4$ | 0.67 cell/day | [1] |
| $\mu_{T_0}$ | $6.7 \times 10^{-7}$ cell/day | [28] | $\theta_1$ | 0.8 cell/day | [21] |
| $\mu_{T_1}$ | 0.3333 cell/day | [31] | $\theta_2$ | 0.6 cell/day | [21] |
| $\mu_{T_2}$ | 0.3333 cell/day | [31] | $\chi_1$ | 1.5 cell/day | [21] |
| $\mu_{M_0}$ | 0.011 cell/day | [21, 31] | $\chi_2$ | 2.0 cell/day | [21] |
| $\mu_{M_1}$ | 0.01 cell/day | [36, 37] | $\beta_1$ | $1.026 \times 10^{-5}$ cell/day | [21, 28] |
| $\mu_{I_M}$ | 0.3264 cell/day | [21, 28] | $\beta_2$ | $1.026 \times 10^{-5}$ cell/day | [21, 28] |
| $\mu_V$ | 0.67 cell/day | [21] | $\tau_1$ | 0.1642 cell/day | [28] |
| $\mu_A$ | 0.332 cell/day | [1, 21] | $\psi$ | 5 cell/day | [21] |
| $\mu_E$ | 0.5 cell/day | [14] | $\sigma$ | 0.5 cell/day | [21] |
| $\mu_{N_K}$ | 0.42 cell/day | [38] | $\omega$ | 20 cell/day | [21] |
| $\mu_{I_{10}}$ | $3.70 \times 10^{-2}$ cell/day | [31] | $\varepsilon$ | 1 cell/day | [21] |
| $\mu_{F_1}$ | 4.9 cell/day | [21] | $\pi$ | 0.3359 cell/day | [28] |
| $\mu_{F_2}$ | 5.16 cell/day | [21] | $\zeta$ | 0.01 cell/day | [21, 32] |

Linearized system 1 about the equilibrium state 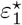, we obtain a Jacobian matrix of the form:

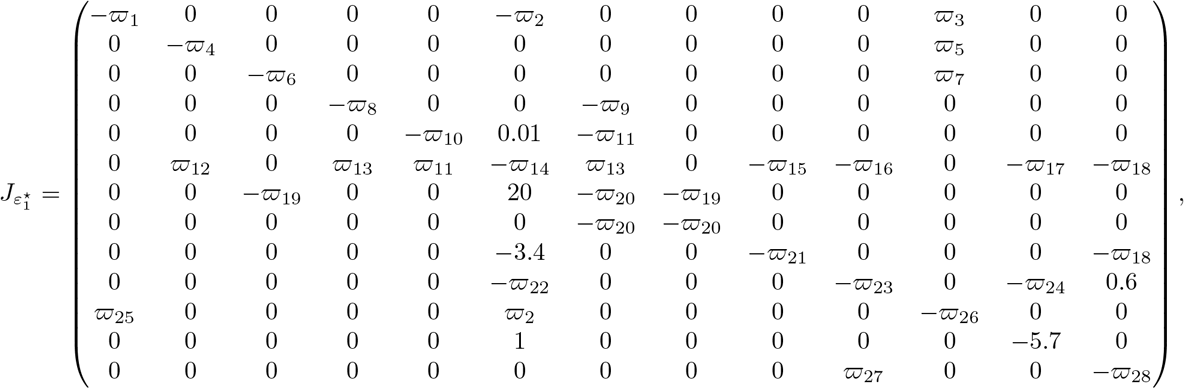

where

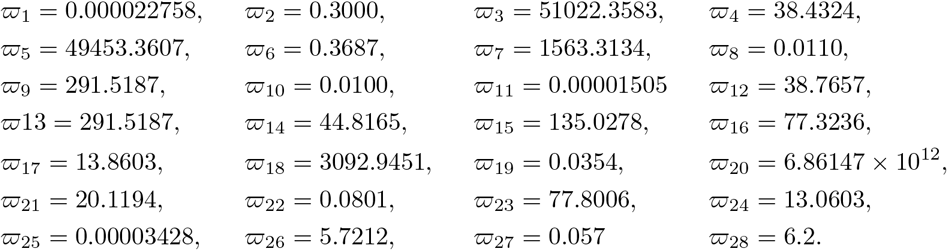

The coexisting equilibrium point 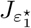 exhibits eigenvalues

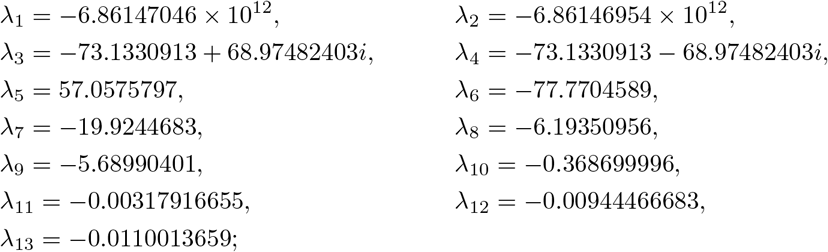

corresponding to an unstable inward spiral. The production of antibodies, effector B cells, naive T cells, and natural killers renders the virus persistence equilibrium state irrelevant. This stabilization promotes stability in the virus-free equilibrium, facilitating viral clearance.

Case II: For the second scenario where *V ≠* 0, 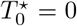, *E*^⋆^ = 0,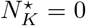, we obtain a boundary steady state of the form:

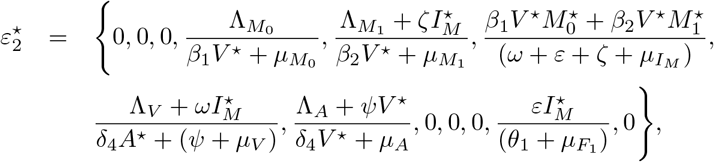

Considering the initial conditions for the state variables and parameter values in Table 1, we have

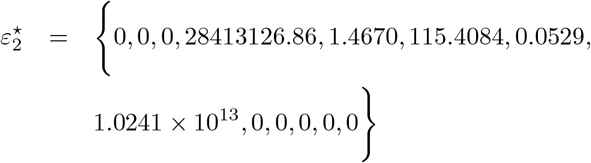

The eigenvalues of 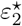 are:

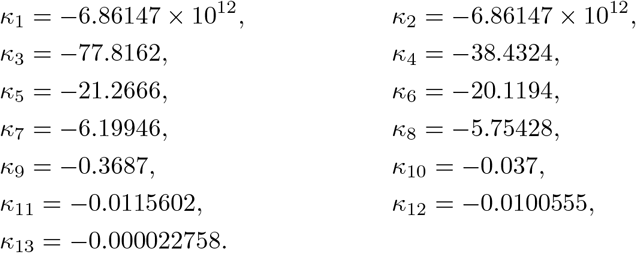

This indicates that 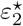 is a saddle-type critical point. Therefore, around this point, the system exhibits stable behavior. This equilibrium is considered a chronic equilibrium because there are no T cells, natural killer cells, effector B cells, or cytokines that can clear the virus. Thus, the presence of antibodies alone is not sufficient to clear the viral infection.

Case III: For the third scenario where *V* ^⋆^ *≠* 0, 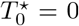, *E*^⋆^ = 0, 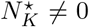, we obtain a boundary steady state of the form:

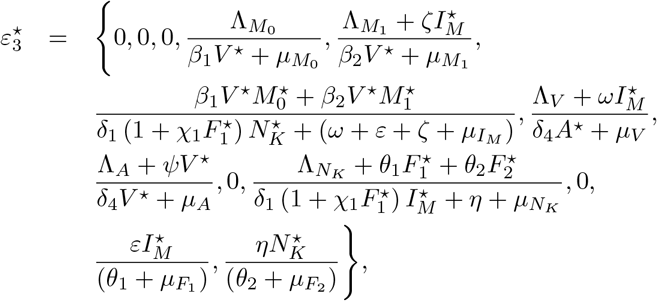

Considering the initial conditions for the state variables and parameter values in Table 1, we have

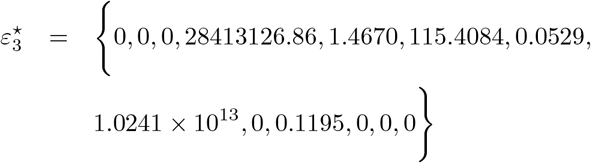

The eigenvalues of 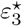 are:

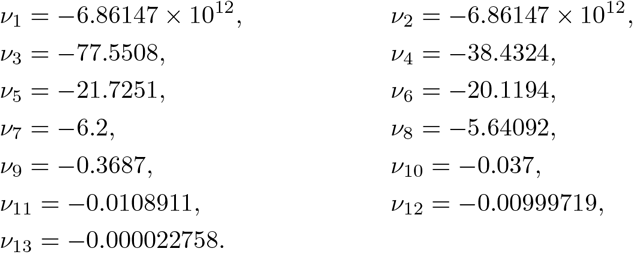

Regarding the equilibrium 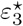, the eigenvalues indicate that it is a saddle point, and the system is stable around 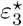. As T cells, effector B cells, and other cytokines such as interferon alpha, beta, and gamma are absent at this equilibrium, there is no chance of viral clearance. This implies that the presence of antibodies and natural killer cells alone is not sufficient to destabilize the virus-persistence equilibrium required for viral clearance.

Case IV: For the fourth scenario where *V* ^⋆^ *≠* 0, 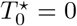, *E*^⋆^ *≠* 0, 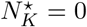, we obtain a boundary steady state of the form:

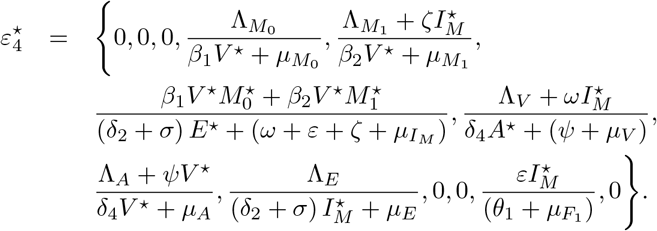

Considering the initial conditions for the state variables and parameter values in Table 1, we have

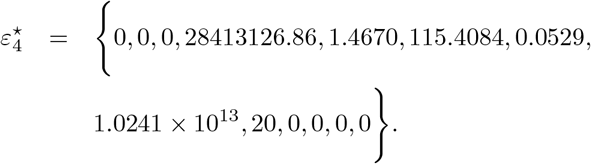

The eigenvalues of 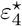 are:

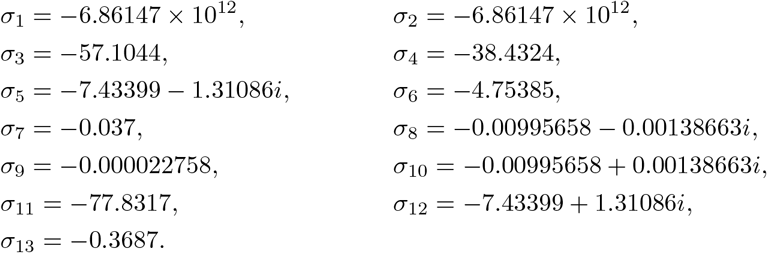

Corresponding to the equilibrium point 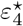, the eigenvalues indicate it is a stable inward spiral, and the system remains stable around this point. In this equilibrium state, the absence of T cells, natural killer cells, and other cytokines such as interferon alpha, beta, and gamma means there is no possibility of viral clearance. This indicates that the presence of antibodies and effector B cells alone cannot disrupt the virus-persistence equilibrium needed for viral clearance.

Case V: For the fifth scenario where *V* ^⋆^ *≠* 0, 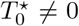, *E*^⋆^ = 0, 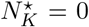, we obtain a boundary steady state of the form:

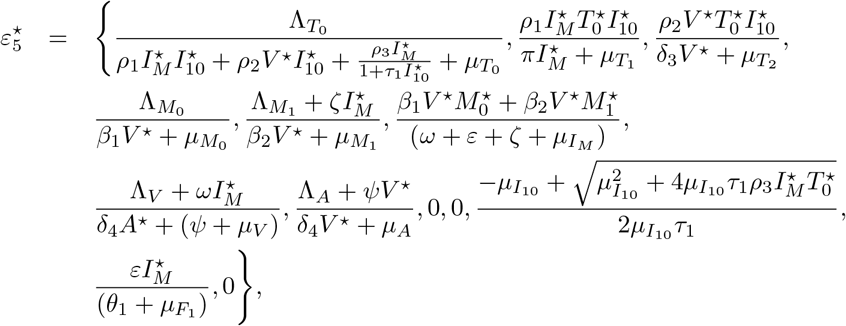

Given the initial conditions for the state variables and parameter values in Table 1, we have

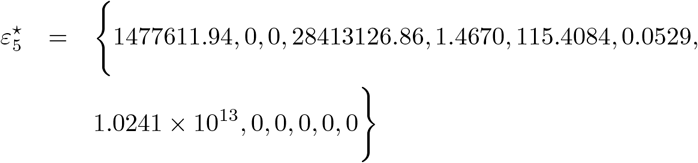

As corresponding to the equilibrium 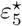, the eigenvalues are

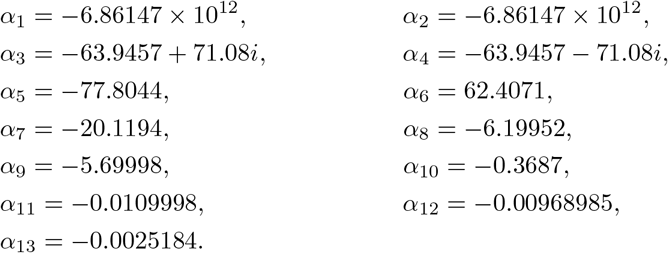

The equilibrium 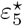 is an unstable saddle point. Biologically, boosting the production of T cells, aided by antibodies, is sufficient to disrupt the virus persistence equilibrium and stabilize the virus-free equilibrium. Consequently, the virus is cleared.

Case VI: For the sixth scenario where *V* ^⋆^≠ 0, 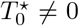, *E*^⋆^≠ 0, 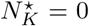, we obtain a boundary steady state of the form:

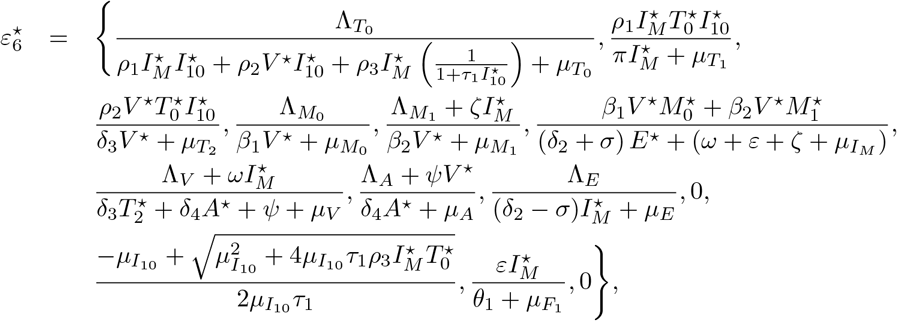

The equilibrium state 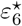 will exists provided that *δ*_2_ *> σ*. Given the initial conditions for the state variables and parameter values in Table 1, we have

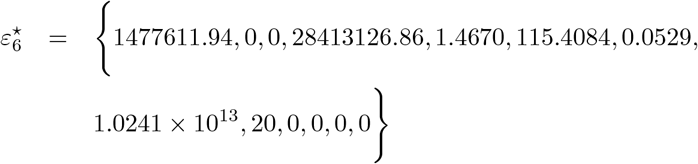

The eigenvalues of 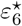 are

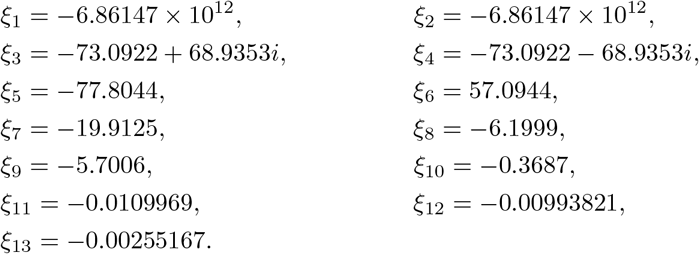

Referring to the eigenvalues of 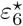, this equilibrium is an unstable node. The presence of naive T cells and effector B cells, along with antibody support, is adequate to destabilize the virus persistence equilibrium and thereby facilitate virus clearance.

Case VII: For the seventh scenario where *V* ^⋆^ ≠ 0, 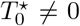, *E*^⋆^ = 0, 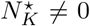, we obtain a boundary steady state of the form:

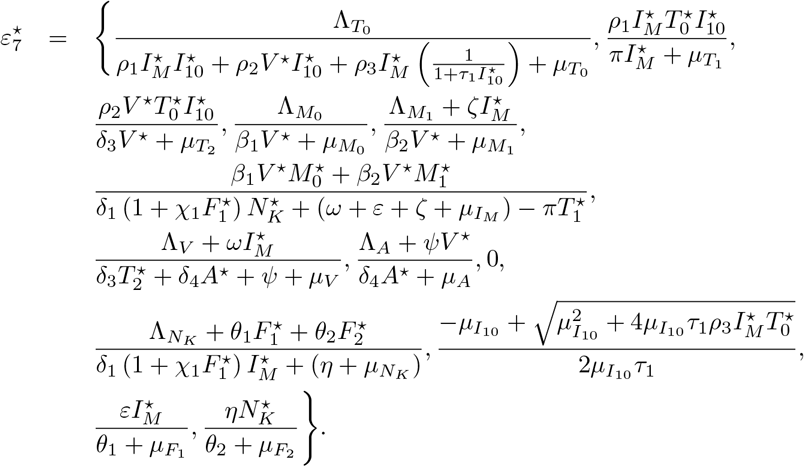

The equilibrium 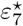 will exists if 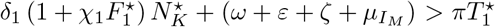. Given the initial conditions for the state variables and parameter values in Table 1, we have

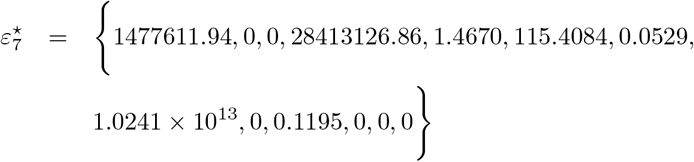

The eigenvalues of the Jacobian matrix 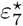 are

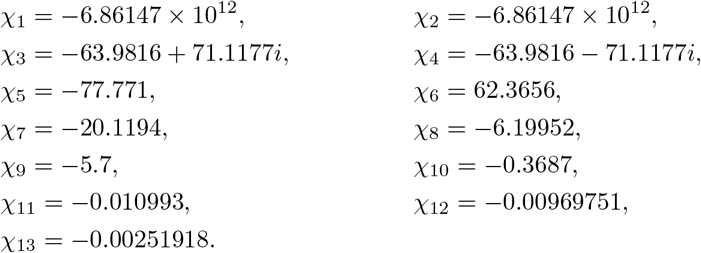

The eigenvalues of 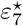 indicate that this boundary steady state is an unstable spiral node. The presence of naive T cells and natural killer cells, along with antibody support, is sufficient to destabilize the virus persistence equilibrium and consequently lead to virus clearance.

Case VIII: For the eighth scenario where *V* ^⋆^ *≠* 0, 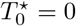, *E*^⋆^ *≠* 0, 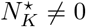, we obtain a boundary steady state of the form:

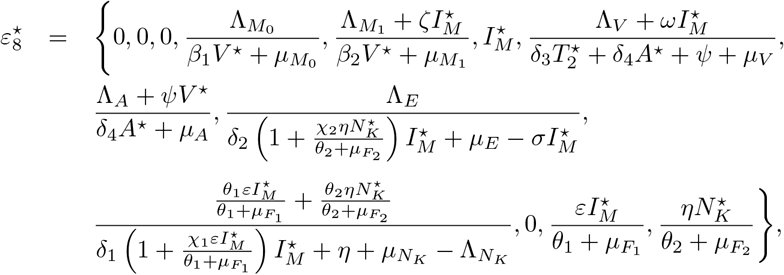

Where 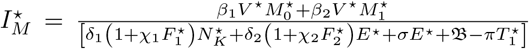, and 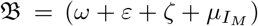. This equilib-rium 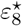 will exists if 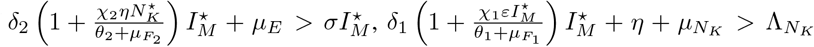 and 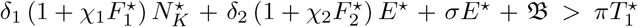. Given the initial conditions for the state variables and parameter values in Table 1, we obtain

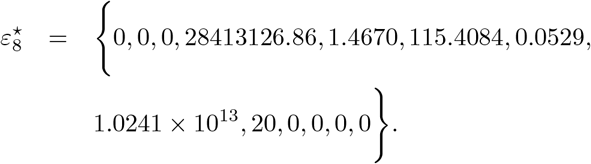

The equilibrium point 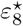 has eigenvalues

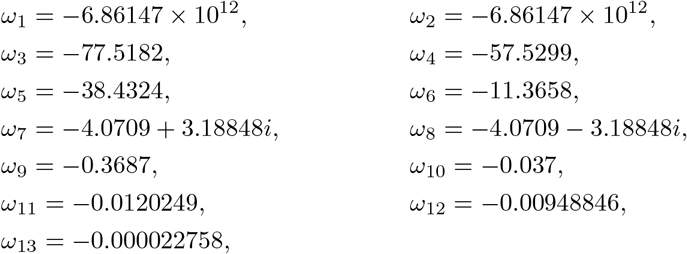

indicating that this boundary steady state is a stable saddle node. The absence of naive T cells results in a stable virus persistence equilibrium. Despite the production of antibodies and effector B cells, they are insufficient to disrupt the virus persistence equilibrium. This underscores the crucial role that T cells play in clearing hepatitis B virus infection.

## 6 Numerical Simulations and Sensitivity Analysis

In this section, we present the numerical simulations and sensitivity analysis of the proposed model. We utilize values provided in Table 1 for the immune system response to the co-existence dynamics of HBV and liver cancer. To evaluate the usefulness and functionality of system (1) numerically, we use a set of reasonably approximated parameter values derived from published HBV article [1]. The following initial conditions: *T*_0_(0) = 5 *×* 10^5^, *T*_1_(0) = 0, *T*_2_(0) = 0, *M*_0_(0) = 4 *×* 10^5^, *M*_1_(0) = 4 *×* 10^5^, *I*_*M*_ (0) = 0, *V* (0) = 300, *A*(0) = 0, *E*(0) = 0, *N*_*k*_(0) = 0, *I*_10_(0) = 0, *F*_1_(0) = 0, and *F*_2_(0) = 0 and the parameters used in the numerical simulations are in Table 1.

### 6.1 Numerical Results at Virus-free Equilibrium State

To showcase the various types of dynamical behaviors that model 1 can exhibit in different parameter regimes, we solve this system numerically using the baseline parameter values provided in Table 1. The results are displayed in Figures 2, 3, and 4.

**Figure 2:**
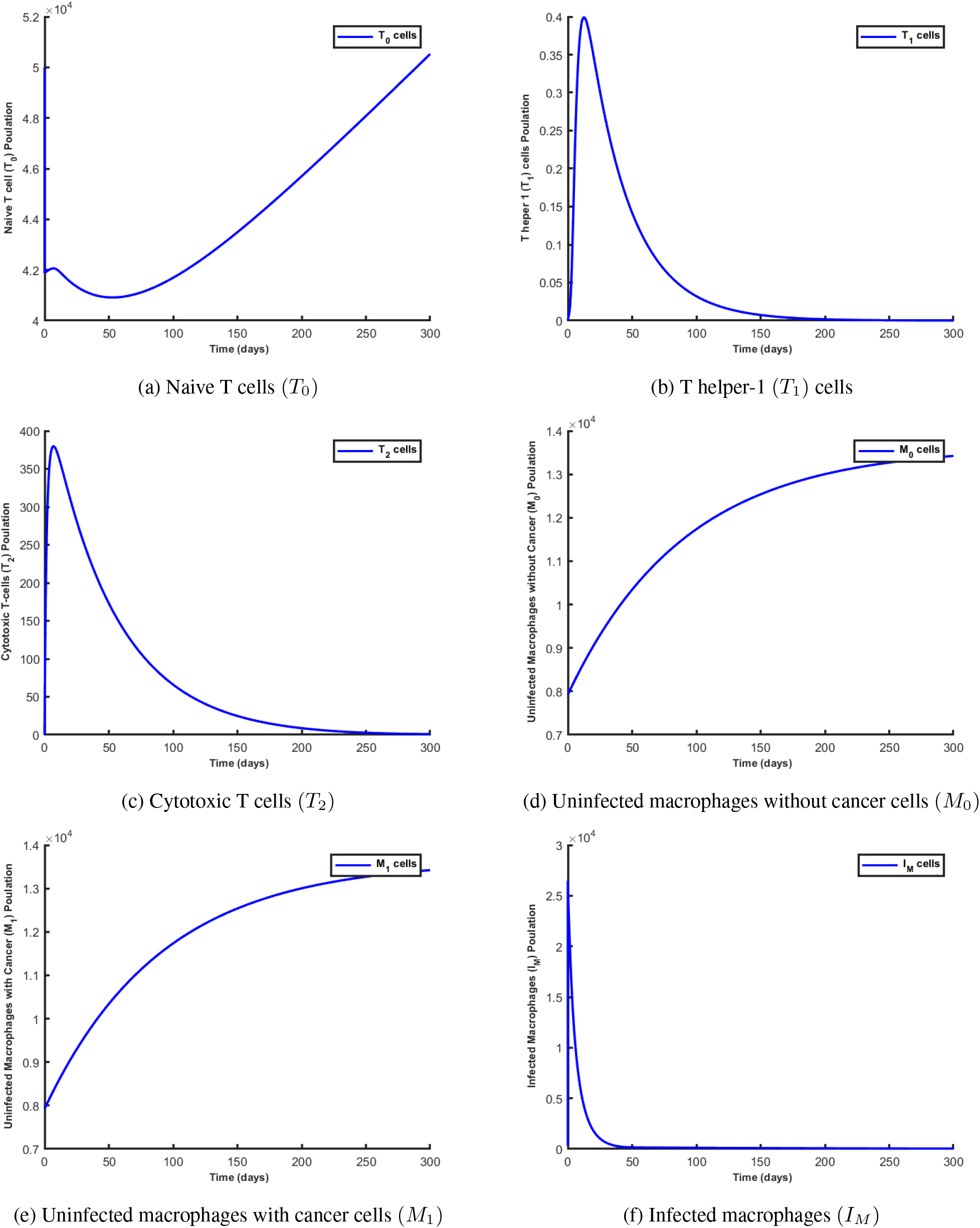
Simulation results showing the behaviour of the state variables at the virus-free equilibrium state.

**Figure 3:**
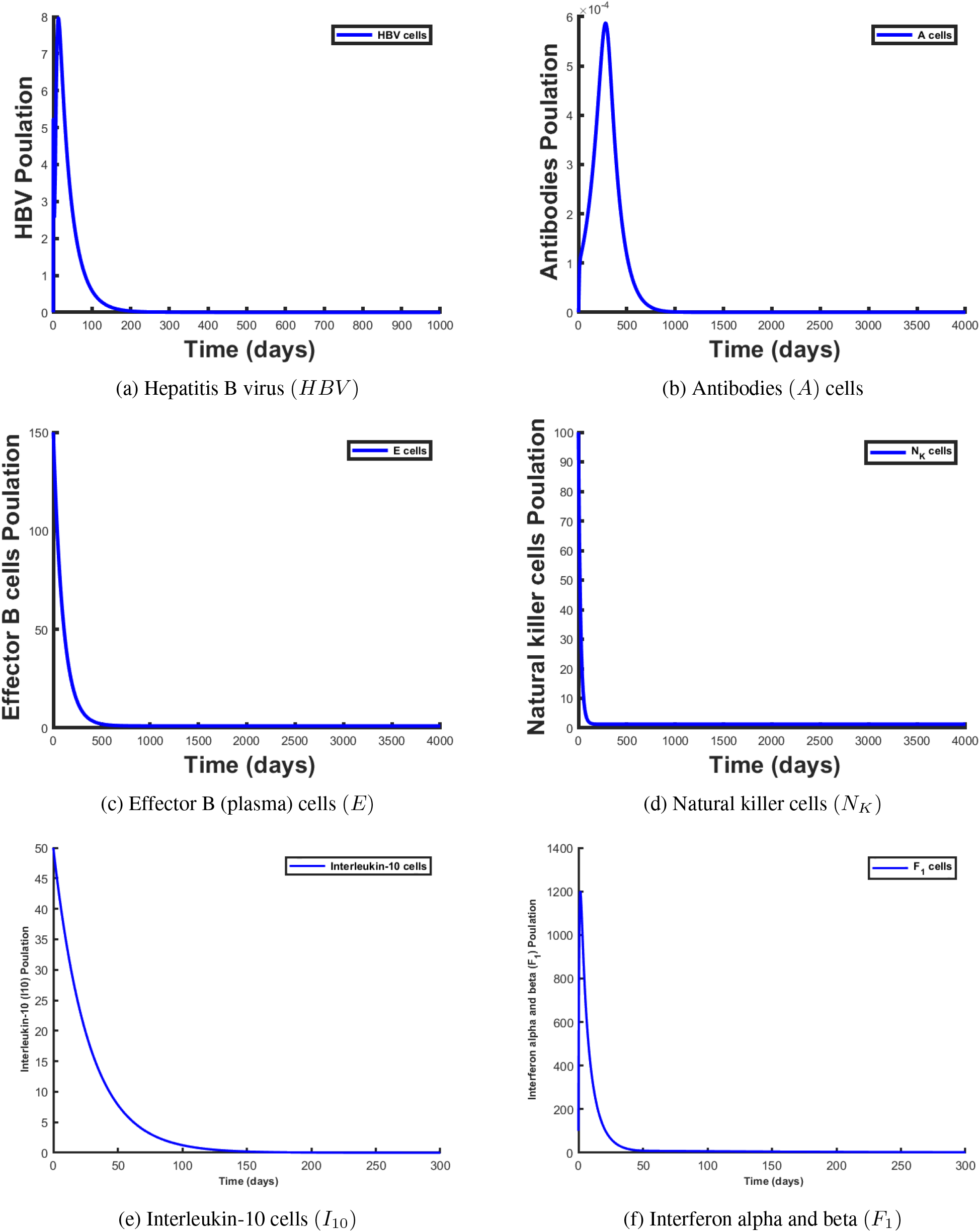
Simulation results showing the behaviour of the state variables at the virus-free equilibrium state.

**Figure 4:**
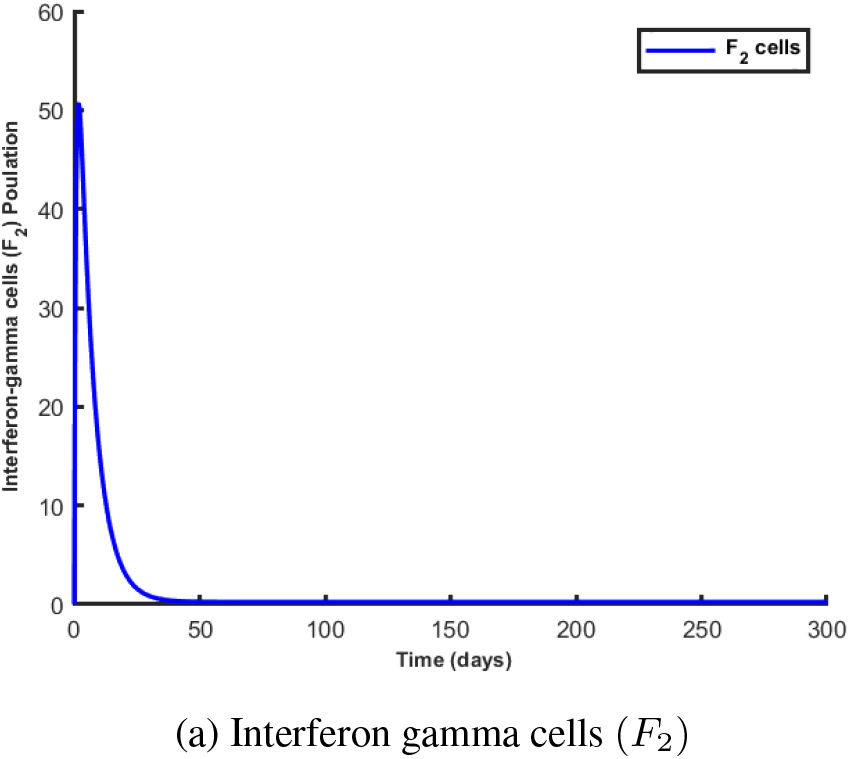
Simulation results showing the behaviour of the state variables at the virus-free equilibrium state.

Figures 2, 3, and 4 depict the immune response dynamics when there is no hepatitis B virus in the liver cell population. In this scenario, the initial viral growth leads to an increase in the numbers of antibodies (*A*), effector

B cells (*E*), natural killer cells (*N*_*K*_), cytotoxic T cells (*T*_2_), and both types of interferons. This results in the successful clearance of the HBV infection, after which type-1 interferons are also eliminated, and the system reaches a stable virus-free steady state. The absence of infected macrophages suggests that hepatitis B virus is not being transmitted, which consequently means that naive T cells will not engage in any lytic activity. Thus, the the T helper-1, cytotoxic T cells, antibodies, effector B cell, natural killer, interleukin-10 and interferon-gamma populations turns to zero as illustrated in Figures 2, 3 and 4 respectively.

### 6.2 Numerical Results at Virus Persistence Equilibrium State

Figures 5, 6, and 7 illustrate the behavior profiles of the state variables of model 1 after infection with a few hepatitis B viruses, at the virus persistence equilibrium. From Figure 5(a), we observe the dynamics of naive T cells being fully produced with a total density of approximately 5 *×* 10^5^. However, when HBV is introduced into the liver cells, the dynamics of naive T cells change. The presence of HBV, whose dynamics are depicted in Figure 6(a), causes a decrease in the total density of naive T cells. This decrease is due to the differentiation process they undergo to produce T-helper 1 (*T*_1_), cytotoxic T cells (*T*_2_), and interleukin-10 (*I*_10_) cells.

**Figure 5:**
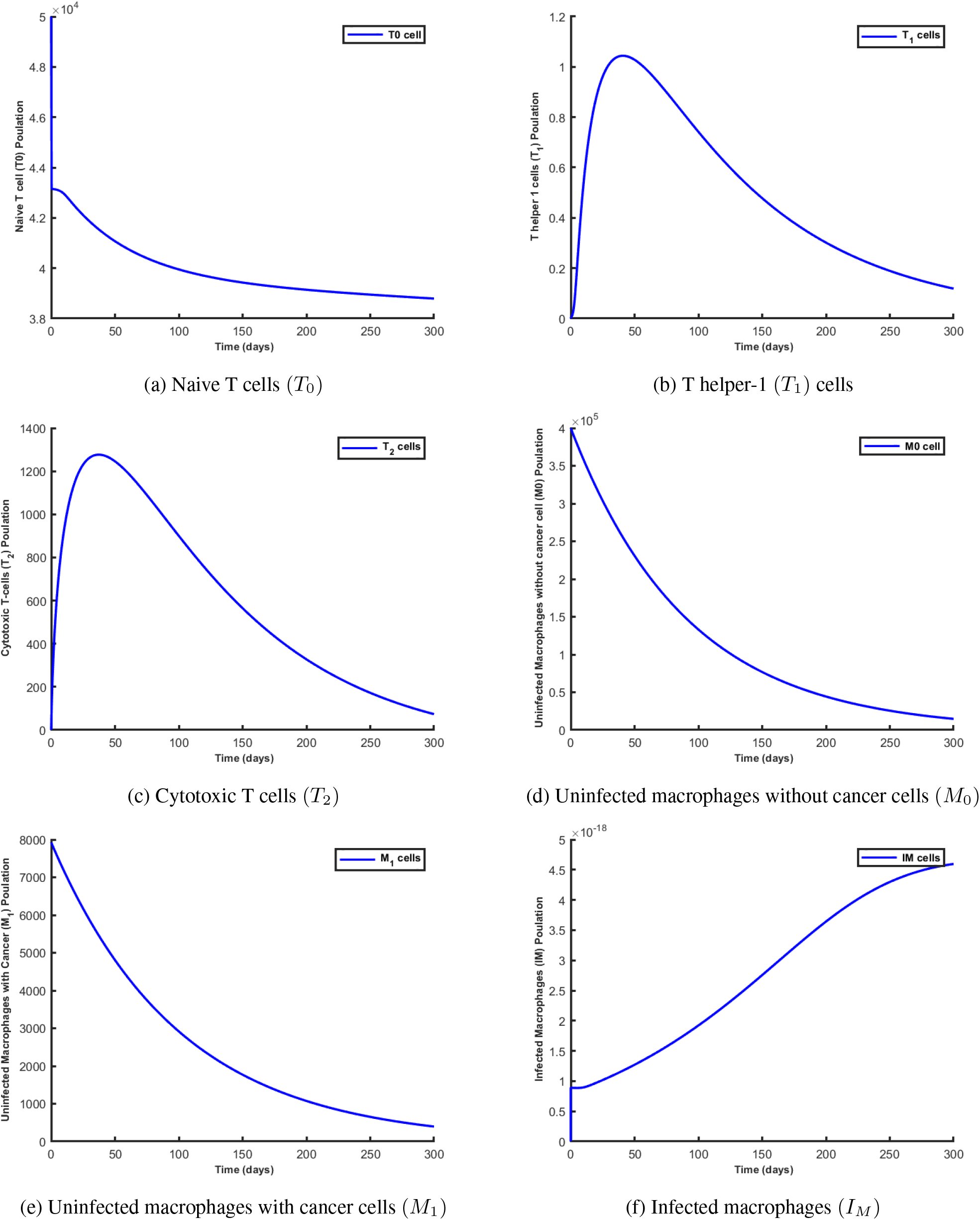
Simulation results showing the behaviour of the state variables at the virus persistence equilibrium state.

**Figure 6:**
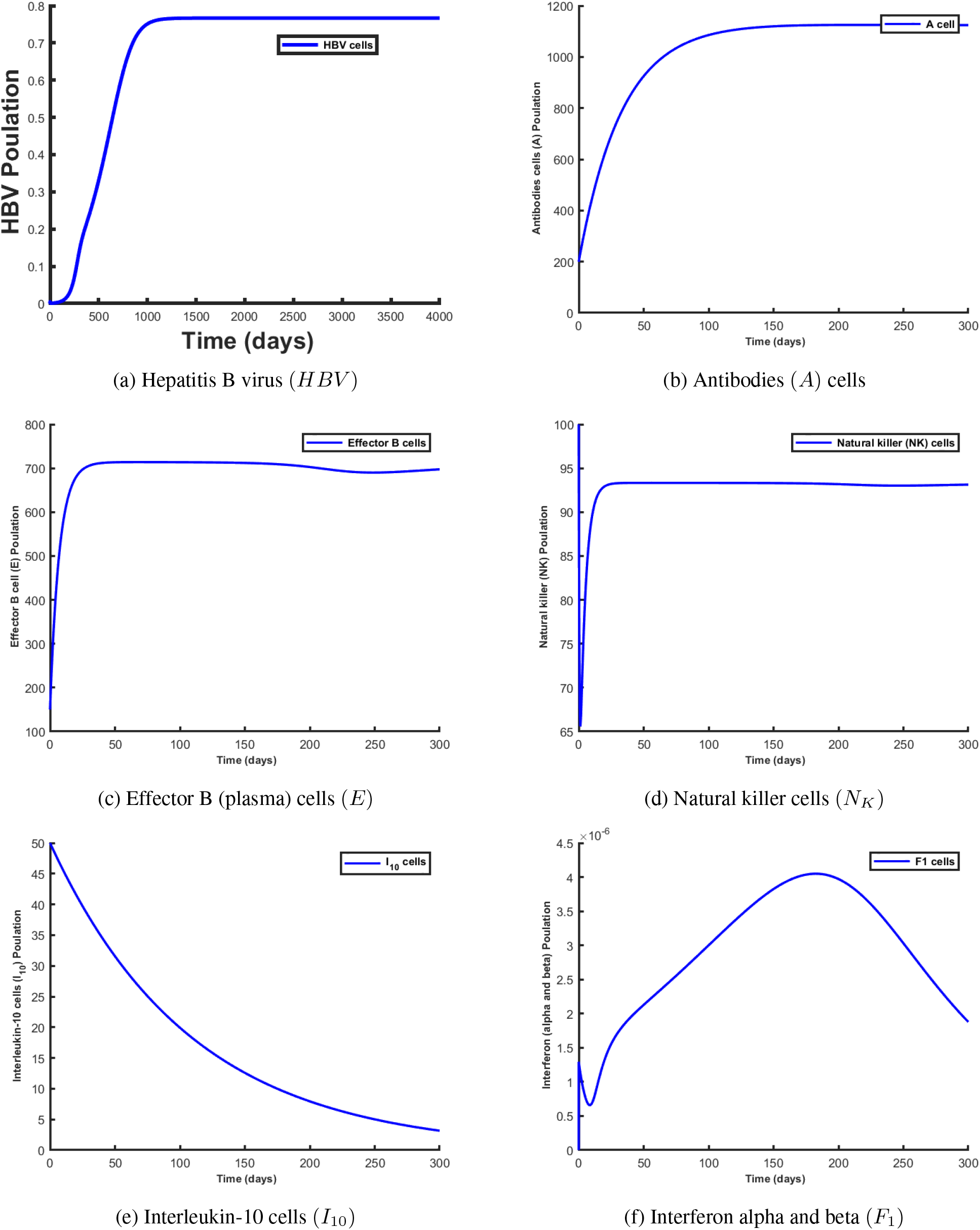
Simulation results showing the behaviour of the state variables at the virus persistence equilibrium state.

**Figure 7:**
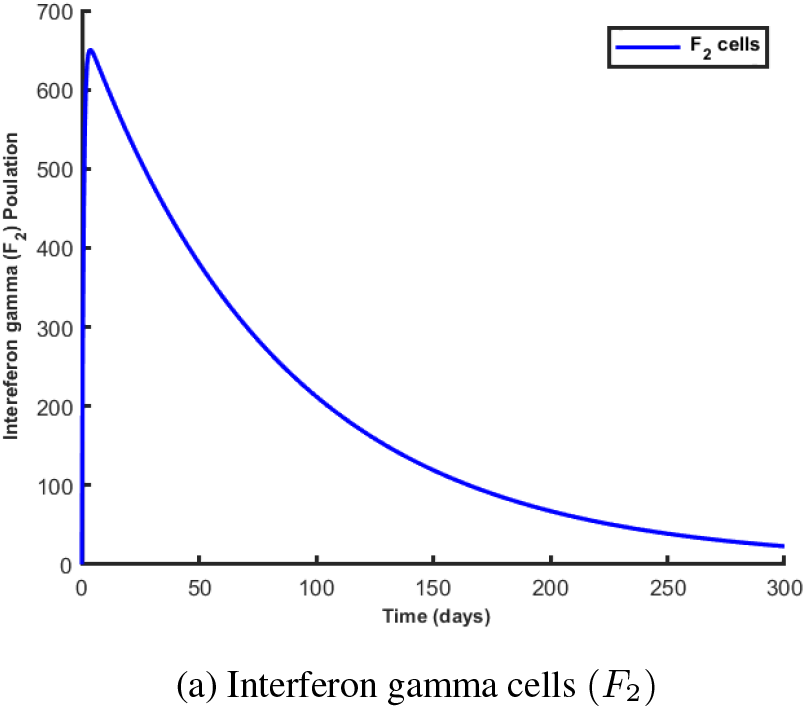
Simulation results showing the behaviour of the state variables at the virus persistence equilibrium state.

We also observed that HBV and naive T cells have a negative correlation, meaning that a higher and more robust naive T cell density leads to a decrease in the HBV load. Consequently, with a constantly increasing HBV load, the rate at which naive T cells decrease accelerates, accompanied by successful production of both pro-inflammatory and anti-inflammatory cells, as shown in Figures 6(e), 6(f), and 7. From Figures 5(b) and 5(c), we examine the dynamics of T helper 1 and cytotoxic T cells, respectively. We notice an initial increase in both cell types due to naive T cell differentiation.

However, we observed that cytotoxic T cells decrease immediately after they are produced within the shortest possible time due to the presence of HBV, which constantly inhibits their production. T helper 1 cells, on the other hand, decrease as a result of lytic activity caused by the replication rate of hepatitis B virus inside macrophages, impacting the overall rate of disease progression. Figures 5(d) and 5(e) show that uninfected macrophages without cancer cells and uninfected macrophages with cancer cells start with an initial density of approximately 4 *×* 10^5^ and 7925, respectively, which decreases over time. Their decreasing nature is attributed to the increasing HBV load.

As the replication rate of hepatitis B virus increases, more macrophages are required at the infection site to engulf the virus, as illustrated in Figure 5(f). This activity decreases the density of the uninfected macrophage population. Consequently, the number of infected macrophages increases as more uninfected macrophages engulf the virus and become infected. However, the rise in infected macrophages stabilizes once they become activated, as shown in Figure 5(f). Furthermore, the density of infected macrophages, which positively correlates with interferon gamma, interferon alpha and beta cytokines, antibodies, effector B cells, and natural killer cells, is illustrated in Figures 6(b), 6(c), and 6(d). As the concentration of interferon gamma increases, as shown in Figure 6(f), it triggers the activation of more infected macrophages, which in turn increases the population of uninfected macrophages. Thus, Figures 5, 6, and 7 depict the dynamics when the endemic steady state is feasible and stable. It can be observed that the initial viral growth is suppressed by the combined effects of different branches of the immune system.

### 6.3 Switching Time

The results of HBV infection differ significantly among individuals, highlighting the importance of host genetic factors in determining susceptibility to HBV persistence and the progression of liver damage to cirrhosis and HCC. An effective antiviral response, primarily mediated by *CD*4^+^ and *CD*8^+^ T-cells, natural killer cells, and monocytes, can lead to immune-controlled HBV replication (functional cure). Conversely, in children and adults with weakened immune systems, active viral replication may persist.

In its primary function, IL-10 acts as an immunosuppressive cytokine by inhibiting T-cell proliferation and the functions of antigen-presenting cells (APCs), as well as by modulating the synthesis of cytokines and chemokines [39]. Recently, a subset of IL-10 producing B-cells, known as regulatory B-cells (Bregs), has been shown to regulate HBV-specific *CD*8^+^ T-cell immunity [40, 41]. Down-regulation of IL-10 restores the function of exhausted HBV-specific *CD*8^+^ T-cells [40]. However, IL-10 may significantly impact the antiviral immune response, as it inhibits the production of pro-inflammatory cytokines such as IFN-*γ*, TNF-*α*, IL-1*β*, and IL-6. In chronic HBV infection, both the number of regulatory T-cells and the levels of inhibitory interleukin-10 (IL-10) and transforming growth factor beta (TGF-*β*) increase, leading to HBV-specific *CD*8^+^ T-cell exhaustion and making viral eradication from the liver impossible [42]. Thus, IL-10 expression is elevated during several chronic viral infections, serving as a viral strategy to down-regulate the host immune response and allow viral persistence in the host [43, 44, 45, 46, 47].

Over the past decade, numerous researchers have attempted to use mathematical models to predict when acute hepatitis B infection transitions to chronic infection and eventually to liver cancer; a period they have termed the “switching time.” We use system 1 to predict when T helper-1 (*T*_1_) cells will outnumber cytotoxic T cells (*T*_2_). The goal is to identify the conditions that lead an individual to progress from the acute stage to the chronic stage as the disease progresses. The period during which T helper-1 (*T*_1_) cells surpass cytotoxic T cells (*T*_2_) is considered crucial for an individual’s progression from the acute infection to the chronic stage. In the initial phase of the virus within infected macrophages, the virus tends to remain dormant while continuing to replicate.

However, within the infected macrophages, there is a battle between the immune response and the hepatitis B virus. For the virus infection to reach the chronic stage, there must be a corresponding increase in viral load. Additionally, due to the complex nature of the hepatitis B virus, a higher viral load poses a significant threat to the host, potentially resulting in chronic disease. Therefore, the concept of switching time aims to identify the minimum time and factors that allow T helper-1 (*T*_1_) cells to quickly overcome cytotoxic T cells (*T*_2_), enabling infected macrophages to turn into acute macrophages for the immune response to take effect. Logically, a shorter switching time should correlate with a lower hepatitis B viral load. In our model, we observed that the parameters *ρ*_1_, *ρ*_2_, and *ρ*_3_ significantly impact the manipulation of switching time and viral load. These parameters are involved in the activities of interferon-alpha and beta, as well as interferon-gamma, and the production of pro-inflammatory cytokines such as IFN-*γ*, TNF-*α*, IL-1*β*, and IL-6. These factors play a significant role in the progression of the disease from the acute stage to chronic and liver cancer stages. Keeping all the other parameter values fixed and varying the value of *ρ*_3_, we obtained the following switching time:

From Figures 8(a) to 8(d), we observed the various switching times for the model. Initially, the density of cytotoxic T cells increases in the first few days, after which it begins to decrease and is surpassed by the gradually increasing density of T helper-1 cells. For a parameter value of *ρ*_3_ = 2.03 *×* 10^−7^, we noticed that it takes over 100 days (approximately 112 days) for T helper-1 cells to surpass cytotoxic T cells. However, when we increase the parameter value of *ρ*_3_ from *ρ*_3_ = 2.03 *×* 10^−7^ to *ρ*_3_ = 2.03 *×* 10^−6^, *ρ*_3_ = 2.03 *×* 10^−5^, and *ρ*_3_ = 2.03 *×* 10^−4^, the switching time increases to 266, 457, and 600 days, respectively. This seems unexpected since one would usually expect that producing more interleukin-10 cytokines for the same viral load would help eliminate the infection. However, these findings support the argument that in HBV infection, ongoing antigen presentation by infected cells and exposure to high antigen levels are associated with *CD*8^+^ T cell exhaustion [48]. Biologically, the continuous production of interleukin-10 cytokines impairs cytotoxic T cells (*T*_2_), thus increasing the switching time. A switching time of approximately 112 days corresponds to the initial period after exposure to a core hepatitis B virus (i.e., the acute stage of HBV infection), which is the appropriate time for the immune system to respond. To further investigate and gain more understanding of the dynamics of HBV load as the switching time increases, we vary the parameter *ω* in relation to its switching time value and plot the hepatitis B virus compartment to observe the changes. From Figure 9, we observed that an increase in switching time corresponds to an increase in the viral load. For an *ω* value of 20.000000203, the density of the hepatitis B virus population is depicted by the blue line in the graph. When the value of *ω* is increased to 20.00000203, 20.0000203, and 20.000203, the density of the hepatitis B virus population is represented by the green, red, and magma lines, respectively. Generally, a lower switching time correlates with a lower hepatitis B viral load. Thus, from the graph, we noticed that a decrease in the value of *ω* from 20.000000203 to 20.0000000203, 20.00000000203, and 20.000000000203, which represents a lower switching time, corresponds to a lower hepatitis B viral load as illustrated by the black, cyan, and yellow dashed lines. This result is not surprising since we did not incorporate a time delay into the dynamics of our naive T cells. The lack of a time delay in the production of naive T cells allows for their continuous exposure to high levels of antigens, resulting in their impairment. Hence, the model predicts that a delay in the differentiation of the naive T cells into T helper 1 cells influences the determination of the switching time.

**Figure 8:**
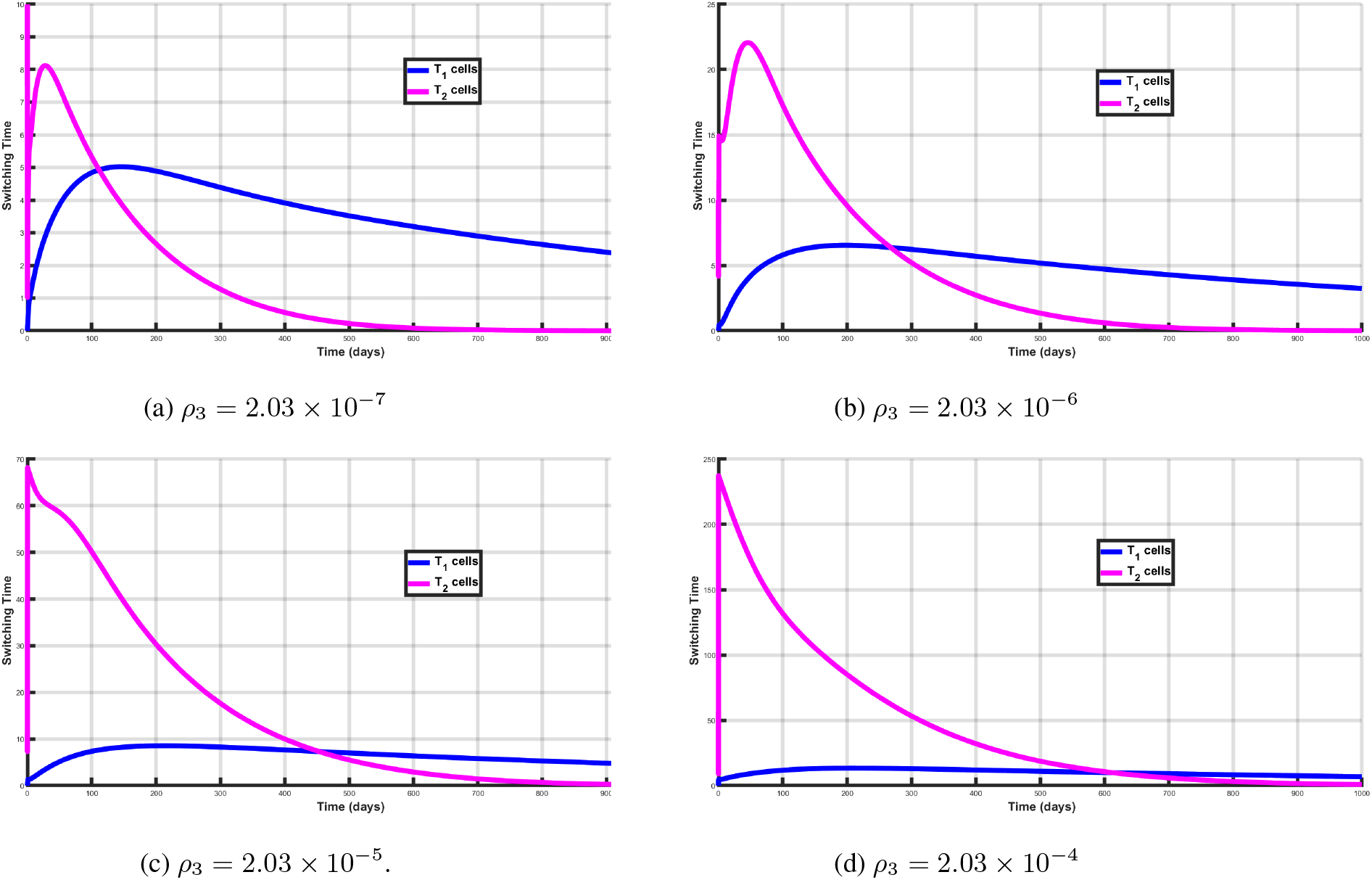
The simulation graph for switching time.

**Figure 9:**
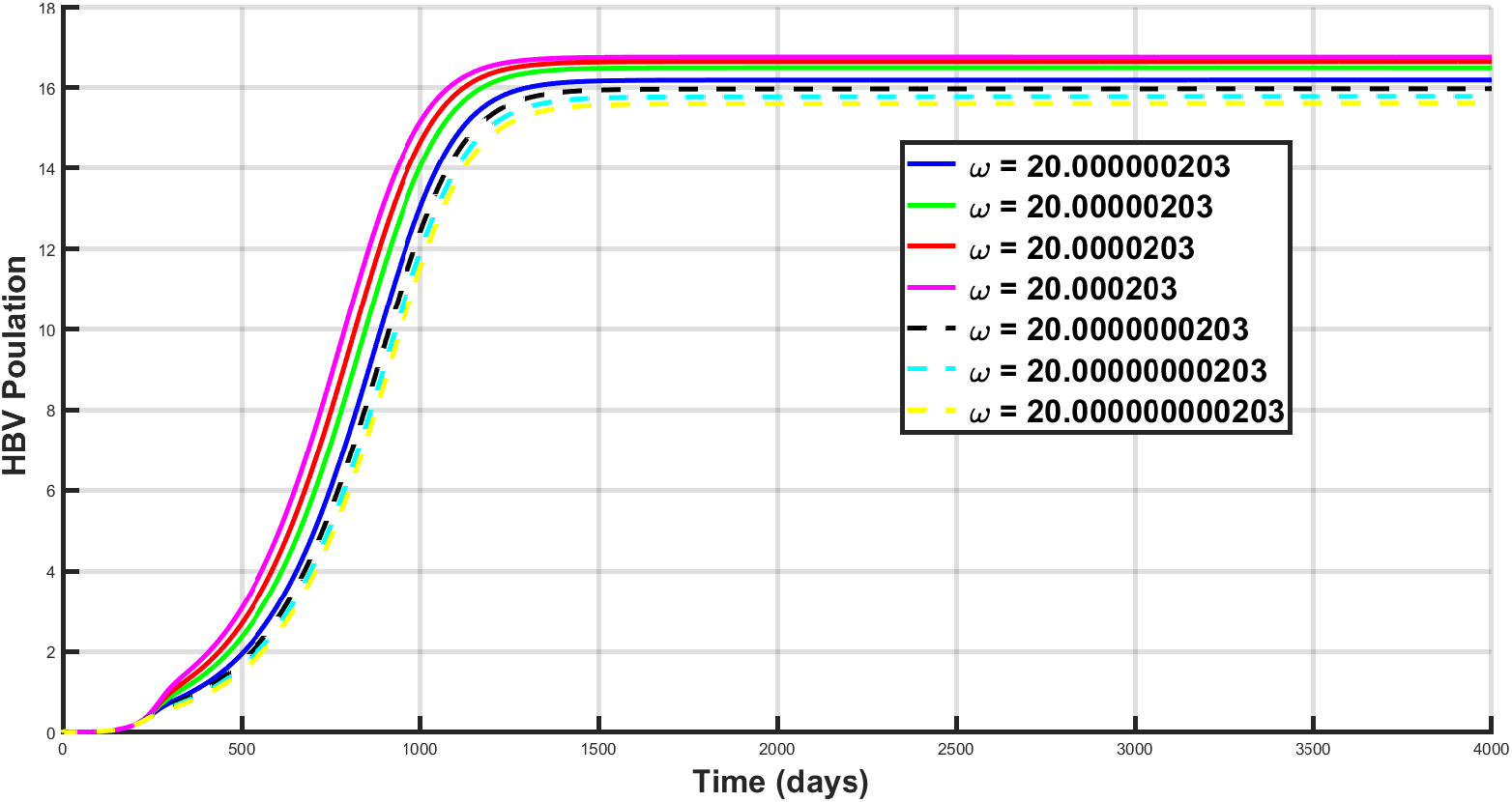
Graph illustrating the numerical solution that depicts the variation in HBV load with different switching times.

### 6.4 Sensitivity Analysis

Due to uncertainties in the collection of data and estimated parameter values, the reproduction number ℝ_0_ is usually affected. We carry out sensitivity analysis to determine the relative importance of each epidemic parameter for transmission and control of both hepatitis B virus and liver cancer diseases and changes in the structure of the model. Latin Hypercube Sampling (LHS) and Partial Rank Correlation Coefficients (PRCC) are employed to identify model parameters that exert the greatest influence on the model, with the reproduction number (ℝ_0_) acting as the dependent function. The main aim of this analysis is to ascertain the effects of parameters on model results. Parameters that are highly sensitive should be estimated with greater precision, as small changes in these parameters can lead to significant variations in the results [49, 50, 51]. Conversely, parameters that are not sensitive require less effort to estimate, since small changes in these parameters do not lead to large variations in the quantity of interest [50]. Parameters with PRCC values greater than positive 0.50 are considered to be highly positively correlated with the dependent function, while those with values less than negative 0.50 are considered to be highly negatively correlated with the dependent function [49, 50, 51]. The parameters included in the PRCC analysis are the interaction rate between *V* and *M*_0_ (*β*_1_), the interaction rate between *V* and *M*_1_ (*β*_2_), the supply rate of uninfected macrophages without cancer 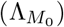, the supply rate of uninfected macrophages with cancer 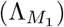, the rate at which infected macrophages develop into liver cancer (*ζ*), the replication rate of new infectious virions (*ω*), the decay rate of uninfected macrophages without cancer 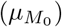, the decay rate of uninfected macrophages with cancer 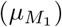, the virus decay rate (*µ*_*V*_), the decay rate of infected macrophages 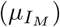, the virus lytic effect rate to release antibodies (*ψ*), and the lytic effect rate of infected macrophages to release interferon alpha and beta cytokines (*ε*). A PRCC analysis was conducted for five different periods; however, the parameters exhibited the same effect on the dependent function across all five periods. Therefore, we chose to present one plot, as shown in Figure 10. The results indicate that the twelve parameters that most significantly impact the response function (ℝ_0_) are *β*_1_, *β*_2_, 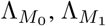, *ζ*, 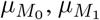, *µ*_*V*_, 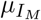, *ψ*, and *ε*. Base on the PRCC values, the parameters *β*_1_, *β*_2_, 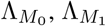, and *ω* positively influence (ℝ_0_), meaning that an increase (decrease) in these parameters will increase (decrease) (ℝ_0_). In contrast, the parameters *ζ*, 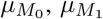, *µ*_*V*_, 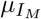, *ψ*, and *ε* negatively influence (ℝ_0_), and an increase in these parameters will decrease (ℝ_0_). The results from PRCC analysis are summarized in Table 2.

**Table 2:**
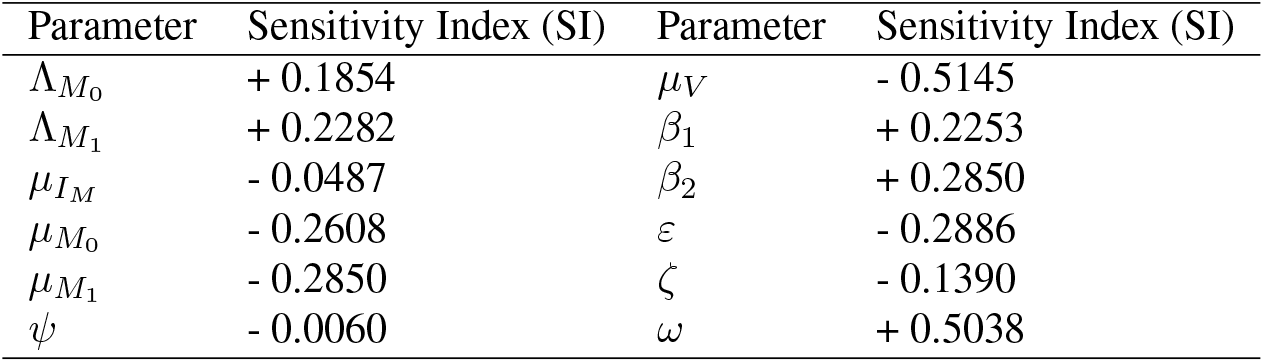
Parameters and their relationship with ℝ_0_.

| Parameter | Sensitivity Index (SI) | Parameter | Sensitivity Index (SI) |
| --- | --- | --- | --- |
| $\Lambda_{M_0}$ | + 0.1854 | $\mu_V$ | - 0.5145 |
| $\Lambda_{M_1}$ | + 0.2282 | $\beta_1$ | + 0.2253 |
| $\mu_{I_M}$ | - 0.0487 | $\beta_2$ | + 0.2850 |
| $\mu_{M_0}$ | - 0.2608 | $\varepsilon$ | - 0.2886 |
| $\mu_{M_1}$ | - 0.2850 | $\zeta$ | - 0.1390 |
| $\psi$ | - 0.0060 | $\omega$ | + 0.5038 |

**Figure 10:**
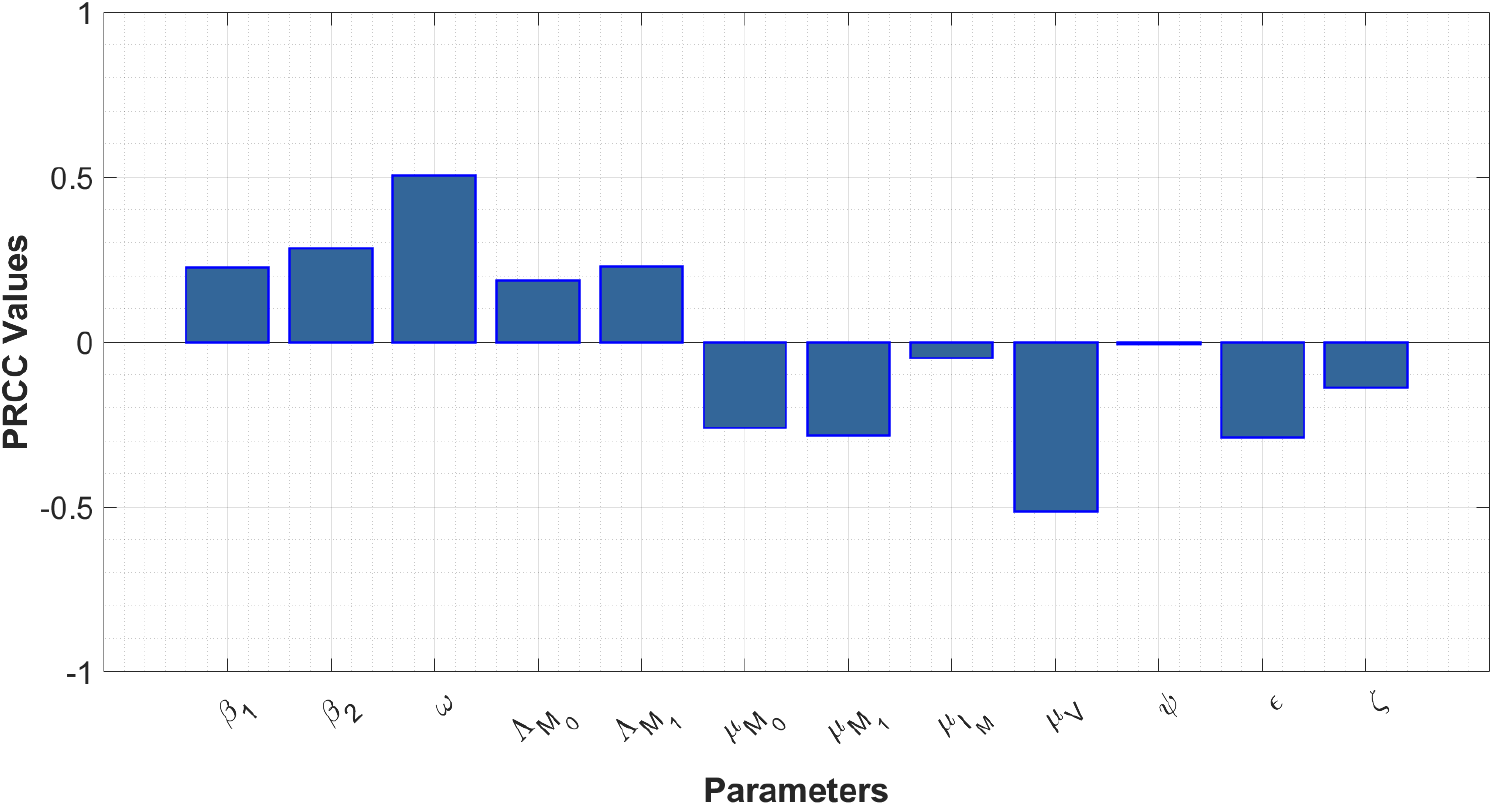
Partial rank correlation coefficients (PRCCs) showing the impact of twelve model parameters on the reproduction number (ℝ_0_) of the model. Parameter values used are as given in Table 1.

## 7 Discussion of results

In this paper, we proposed and examined a new model for HBV infection and its coexistence with liver cancer, with a specific emphasis on the interactions between different branches of the immune system. This model includes the innate immune response, represented by natural killer cells (NK), and the adaptive immune response, represented by effector B cells and naive T cells, which differentiate into T helper-1 cells, cytotoxic T cells, interleukin-10, antibodies, and various cytokines. During infection, cytokines play a crucial role in recruiting both innate and adaptive immune components, boosting their efficacy, and enabling the non-cytolytic clearance of infected cells.

The stability analysis of the steady states demonstrated how different parameters influence the dynamics of the immune response. Some findings, like the destabilization of the virus persistence equilibrium due to the presence of T cells, were intuitively obvious, while others were quite surprising. Naturally, increasing the number of NK cells, the rate at which antibodies clear free virus, the rate at which effector B cells eliminate infected cells, the rate at which cytotoxic T lymphocytes clear infected cells, and the rate at which IFN-*γ* inhibits viral production all contribute to more effective infection clearance, stabilizing the disease-free steady-state solutions. However, we noticed that for a very small or a very large rate of free virus clearance by antibodies, NK cells, and effector B cells, the stability of the endemic steady state is unaffected by how quickly the new antibodies, NK cells, and effector B cells are produced. But whenever the production of naive T cells increases, the virus-persistence equilibrium is destabilized. This result is very astonishing, as one would generally presume that a higher production rate of antibodies, NK cells, and effector B cells would lead to the clearance of the infection rather than the stabilization of a chronic state. This indicates that it is not the distinct production rates of antibodies, NK cells, and effector B cells that lead to viral clearance, but rather the ratio between these rates that determines whether the system maintains a chronic infection, exhibits periodic oscillations, or achieves viral clearance.

Interleukin-10 (IL-10) is a cytokine with anti-inflammatory properties that plays a complex role in the immune response to hepatitis B virus (HBV) infection. IL-10 is primarily recognized for its ability to suppress the immune response. It is well known that IL-10 inhibits the production of pro-inflammatory cytokines such as IL-1, IL-6, TNF-*α*, and interferon-gamma (IFN-*γ*). By regulating these cytokines, IL-10 helps to decrease the overall inflammatory environment in the liver during HBV infection. IL-10 helps maintain a balance between necessary immune activity to control the virus and the need to limit excessive inflammation that can cause liver damage. This balance is crucial because excessive immune responses can lead to conditions like fulminant hepatitis, while inadequate responses can result in chronic infection. During HBV infection, IL-10 can suppress the activity of various immune cells, including T cells, macrophages, and dendritic cells. This suppression can reduce liver inflammation and damage, potentially preventing severe liver injury. Despite the significant role interleukin-10 plays in HBV infection, it can potentially contribute to viral persistence by dampening the immune system’s ability to clear the virus. By inhibiting the immune response, IL-10 may aid in the persistence of HBV. The suppression of cytotoxic T lymphocytes (CTLs), which are crucial for clearing infected cells, can allow the virus to evade the immune system and establish chronic infection as illustrated in Figure 9. Elevated levels of IL-10 have been associated with poor responses to antiviral therapies in HBV-infected individuals. High IL-10 levels can inhibit the effectiveness of the immune response needed for successful treatment.

We observed various switching times for the model. Initially, the density of cytotoxic T cells increases in the first few days, after which it begins to decrease and is surpassed by the gradually increasing density of T helper-1 cells. However, when we increased the production rate of interleukin-10, the switching time increased. This seems unexpected since one would usually expect that producing more interleukin-10 cytokines for the same viral load would help eliminate the infection. Thus, these findings support the argument that in HBV infection, ongoing antigen presentation by infected cells and exposure to high antigen levels are associated with *CD*8^+^ T cell exhaustion. Biologically, the continuous production of interleukin-10 cytokines impairs cytotoxic T cells (*T*_2_), thus increasing the switching time. A switching time of approximately 112 days corresponds to the initial period after exposure to the core hepatitis B virus (i.e., the acute stage of HBV infection), which is the appropriate time for the immune system to respond. However, further production of interleukin-10 beyond this period will lead to immune impairment. The model further showed that under specific conditions (see Figures 8 and 9), the switching time can be adjusted. As illustrated in Figure 9, we observed that a reduction in the value of *ω*, indicating a shorter switching time, corresponds to a lower hepatitis B viral load, as shown by the black, cyan, and yellow dashed lines. This outcome is expected since a time delay was not included in the dynamics of our naive T cells. Consequently, the model suggests that a delay in the differentiation of naive T cells into T helper 1 cells affects the determination of the switching time. Therefore, it is reasonable to state that a positive lifestyle correlates positively with immune system functioning and is crucial in determining whether an acute infection progresses to the chronic stage. The implications of the model align with our expectations; logically, a longer switching time results in a higher viral load and vice versa.

Minimizing the side effects of IL-10 in HBV infection requires carefully managing the immune response to ensure effective viral clearance while preventing excessive immune suppression that could lead to chronic infection or liver damage. Therefore, we recommend developing drugs that selectively inhibit IL-10 activity in specific cell types or tissues to help minimize systemic side effects. Additionally, using antibodies or small molecules to block IL-10 receptors on specific immune cells might reduce the suppression of antiviral responses while preserving the beneficial anti-inflammatory effects in the liver. Combining IL-10 modulation with potent antiviral drugs can lower the viral load, reducing the need for immune suppression. Furthermore, pairing IL-10 inhibitors with other immune modulators that enhance T cell activity or other antiviral immune responses can help balance immune activation and suppression. We have used arbitrary parameter values for simulating the model; it would be better if clinical data sets were used to fit the model. Time delay in the immune response to HBV infection also plays a crucial role in immune response dynamics. Thus, future research can extend this work by incorporating the time-delay factor in the immune response. Another refinement of the model could include different immunotherapy control techniques, such as antibody therapies currently in clinical trials, CAR-T cell therapy, molecular therapy, and exhaustion therapy. These therapies are extensively studied in [12, 21, 52, 53, 54] to find optimal treatment strategies for HBV and liver cancer management.

## Data Availability

All data produced in the present work are contained in the manuscript

## Notes

### Competing Interest Statement

The authors have declared no competing interest.

### Funding Statement

This study did not receive any funding

